# Telemedicine Perspectives of Patients with Non-Dialysis Kidney Disease and Kidney Transplant – A Qualitative Meta-Analysis

**DOI:** 10.1101/2023.12.07.23299612

**Authors:** Christopher D. Manko, Benjamin J. Apple, Alexander R. Chang, Bobbie L. Johannes

## Abstract

**Rationale & Objective:** While the use of telemedicine has increased dramatically across disciplines, patient perspectives on telemedicine related to chronic kidney disease are not well understood. We systematically reviewed qualitative studies on patients with chronic kidney disease to better understand these patients’ perspectives related to telemedicine.

**Study Design:** Qualitative Meta-Analysis

**Setting & Study Populations:** Pre-dialysis chronic kidney disease and kidney transplant patients that used telemedicine.

**Selection Criteria for Studies:** English language studies published in the year 2000 and beyond that investigated patient perspectives in a qualitative manner. Works that were not qualitative or did not focus on provider-patient interactive modes of telemedicine were excluded.

**Data Extraction:** 375 papers were pulled from PubMed, Embase, and Academic Science Premier. After filtering, 8 final papers were selected. These papers were critically appraised for quality and were used in the final analysis.

**Analytical Approach:** We developed a codebook to systematically review each of the selected papers through qualitative meta-analysis.

**Results:** Four primary themes were identified (autonomy, logistics, privacy/confidentiality, and trust) with additional subthemes and further subdivisions to signify positive versus negative experiences. The majority of subthemes and subdivisions (n=9) identified were positively attributed by patients compared to negative attributes (n=6). The subtheme most commonly found was avoiding travel to the hospital, which was identified in all 8 papers. There was substantial variability in the number of papers demonstrating the other subthemes and subdivisions.

**Limitations:** Lack of provider perspectives, non-English studies, and studies published before the year 2000. Papers published after the start of data extraction were also not included.

**Conclusions:** Telemedicine should continue to be offered to patients with kidney disease and kidney transplant patients to facilitate access. Additional research should focus on ways to decrease negative factors experienced by some patients such as difficulty with using the technology.

## Introduction

During the COVID-19 pandemic, the use of telemedicine drastically increased. A large study of over 36 million US individuals, saw an increase from 0.3% in 2019 to 23.6% in 2020 in the ambulatory setting [1]. Reimbursement for telehealth was expanded by the U.S. Department of Health and Human Services (HHS) declaring the COVID-19 pandemic a public health emergency [2]. One group in need of these services is chronic kidney disease (CKD) and transplant patients. CKD can affect multiple systems of the body, and often patients present with comorbid conditions such as hypertension and diabetes [3]. As such, patients need frequent appointments to manage all related conditions. Similarly, transplant patients need close monitoring for the grafted organ and immunosuppressant therapy [4]. Telemedicine offers a solution that may protect patients from potential infections, while also maintaining the necessary continuity of care with providers.

However, challenges with telemedicine have been noted [5]. Prior systematic reviews focused on telehealth and eHealth interventions in dialysis patients have shown conflicting results with potential benefits, however more adequately powered prospective studies are needed [6-8]. As the U.S. HHS ended the Public Health Emergency on 05/11/23, more data on non-dialysis CKD and transplant patient perspectives is needed to guide future decisions about telehealth in this vulnerable population [9]. The goal of this current study is to assess perspectives of non-dialysis CKD and transplant patients in terms of telemedicine.

## Methods

### Qualitative Systematic Review

A systematic review of qualitative literature was performed to assess the perspectives of patients with non-dialysis kidney disease on telemedicine. Prior work on qualitative systematic reviews informed our methodology [10-17]. Dialysis patients were not included as several systematic reviews have been published on this patient population previously [6-8]. The first author (CDM) conducted a literature search on 06/22/2022, using Boolean terms (Supplemental Item 1), from PubMed, Embase, and Academic Science Premier. Inclusion criteria included English language papers published after the year 2000 that were peer-reviewed.

A total of 375 papers were screened and entered into Endnote (Figure 1). After excluding 41 duplicates and irrelevant papers based on primary author review of abstracts, 34 papers were reviewed by the primary author. Of these 34 papers, 8 met the final inclusion/exclusion criteria and were included in this review (Table 1).

**Table 1.**
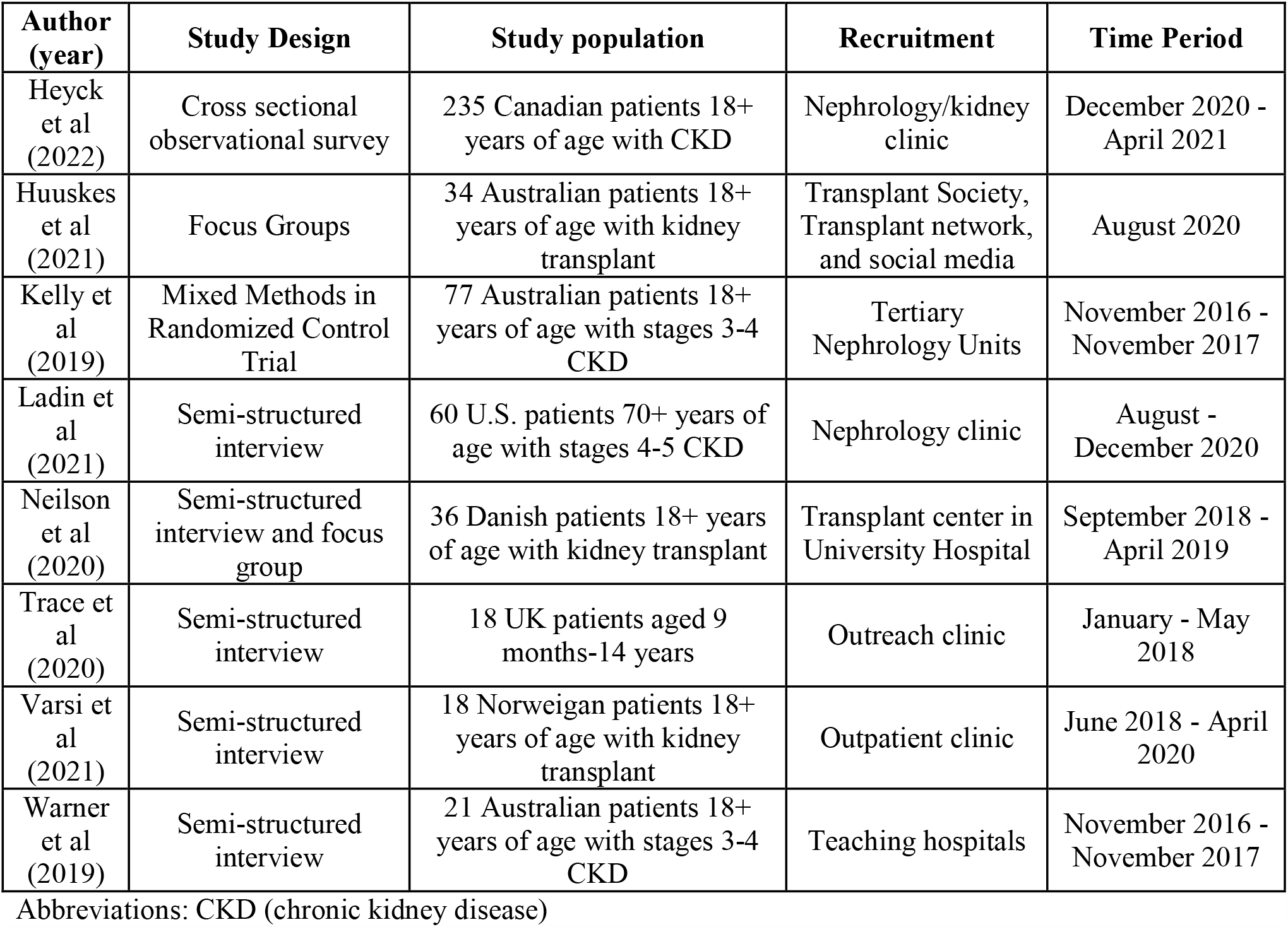
Study characteristics.

**Figure 1.**
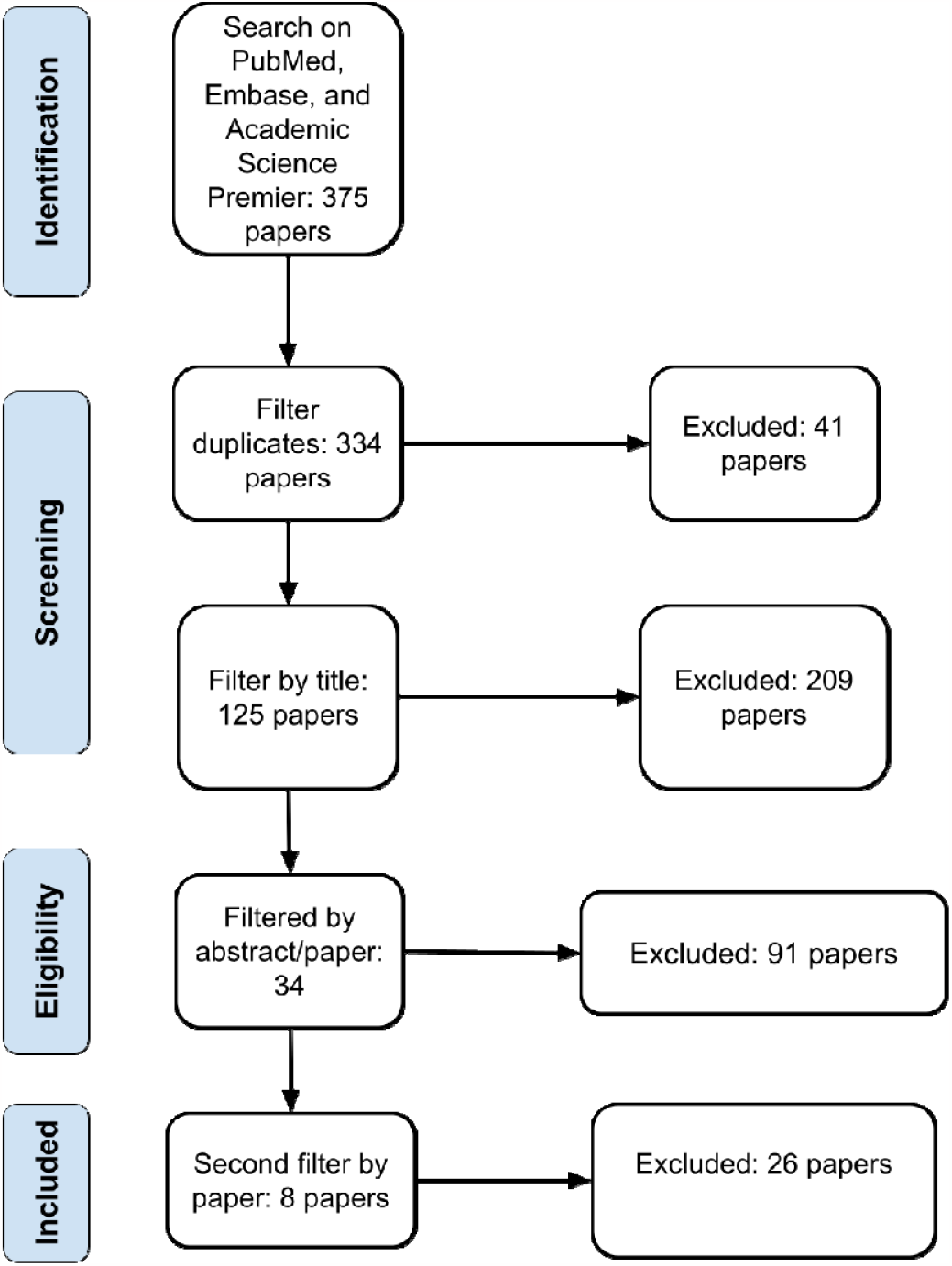
Flowchart of systematic review process

### Qualitative Meta-Analysis

A codebook was created to analyze each paper through an iterative process. The authors independently coded themes and subthemes for 3 of the 8 papers, and then met and consolidated a codebook. Then, 2 of the authors (CDM, ARC) recoded the same 3 papers, reconciled differences in coding though in-person and virtual meetings. The primary author coded the remaining 5 papers and edited the codebook as needed with further consult with the other authors. All coding was performed via NVivo (Release 1.7). Following the completion of coding, a table (Table 2) was created to summarize the quantitative metrics of each code and paper. All 8 papers were assessed for quality via the Crowe Critical Appraisal Tool Form (Supplemental Item 2).

**Table 2.**
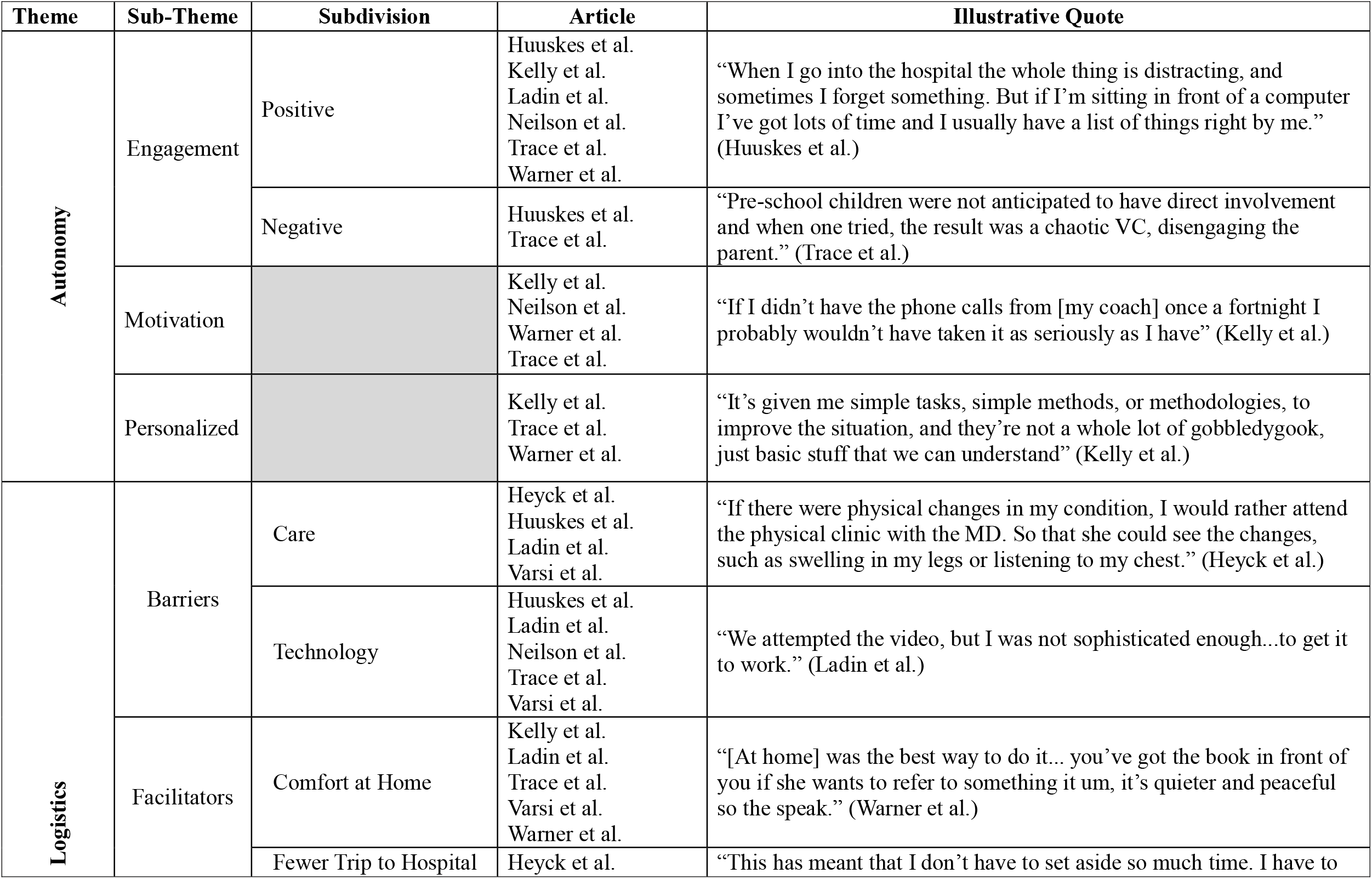
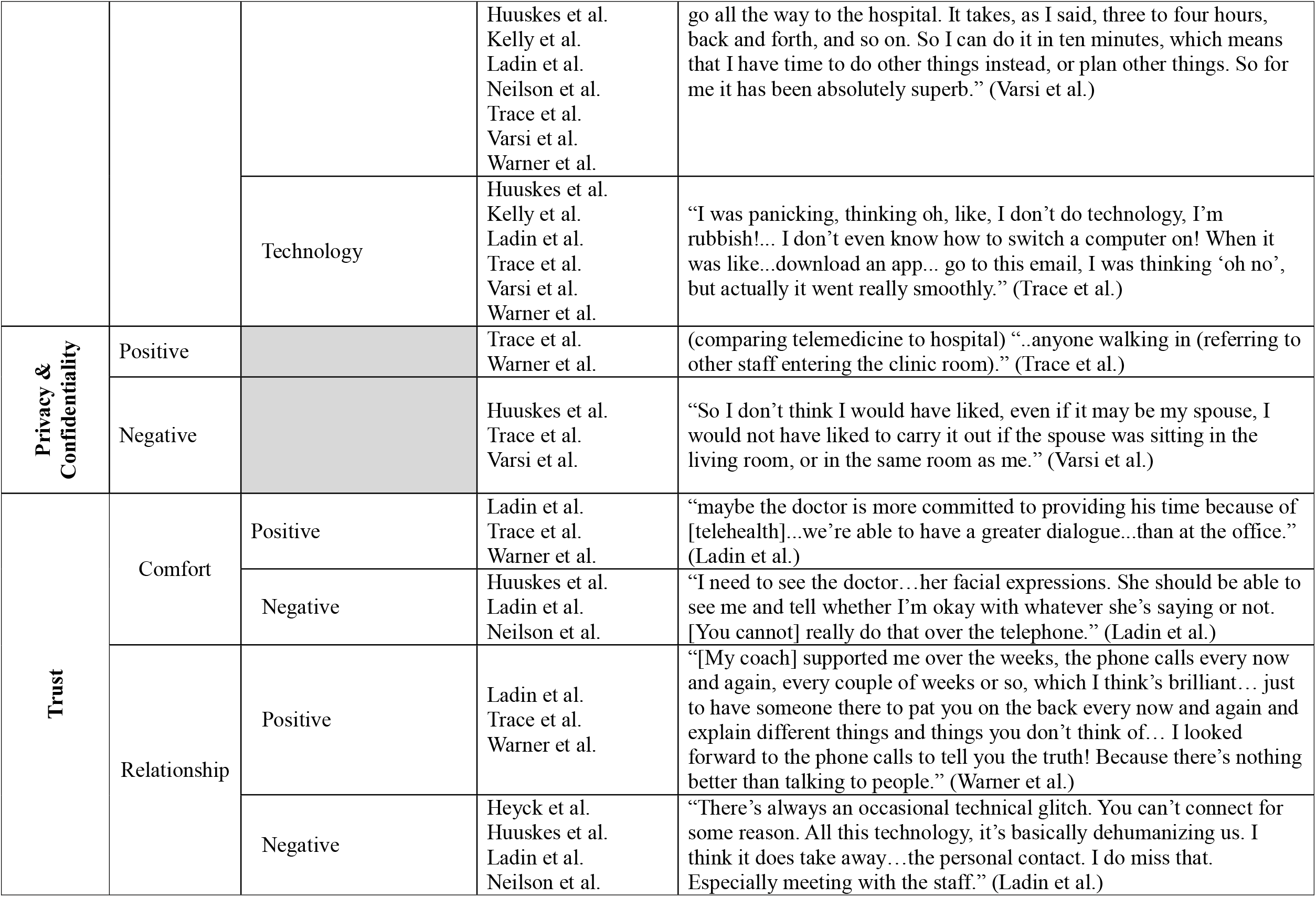
Table of themes, subthemes, and subdivisions of selected studies.

## Results

### Study Characteristics in Systematic Review

Of the 8 studies (N=8) included, 3 focused on transplant patients and 5 focused on non-dialysis CKD patients. 7 focused on adult patients whereas 1 studied a pediatric population. Of the 8 papers, 3 studied dietary interventions. Half were investigated during the COVID-19 pandemic and the other half were pre-pandemic. Characteristics of the studies are shown in Table 1. 3 of the studies utilized telephones as their method of telehealth, 2 studies studied general experiences relating to telemedicine, 2 studies investigated patient experiences with hospital telemedicine applications, and 1 study tested a telehealth app on its participants.

### Themes Identified in Qualitative Analysis

Of the included studies, we identified 4 major themes—autonomy, logistics, privacy and confidentiality, and trust. Table 2 displays illustrative quotes and indicates which themes, subthemes, and subdivisions were present in the papers included in the final analysis.

#### Autonomy

Autonomy had 3 subthemes; engagement (which was further subdivided as positive (n=6) or negative (n=2)), motivation (n=4), and personalized (n=3). Engagement is defined as a patient taking an active part in their healthcare, motivation is a patient’s desire to be engaged in their health and healthcare, and personalized is the care a patient receives being tailored to their specific needs and/or goals.

#### Logistics

There were 2 subthemes within the logistics theme; barrier, which was further contextualized to the subdivisions care (n=4) and technology barriers (n=5), and facilitator which consisted of the subdivisions comfort at home (n=5), fewer trips to hospital (n=8), and technology (n=6). Care is defined as the ability of telehealth to provide adequate care for a patient, whereas technology barriers is defined as issues specific to the telehealth modality which could include obstacles such as lagging, disrupted connections, etc. Comfort at home is defined as a patient’s view of telehealth being beneficial in that care is received at home, fewer trips to the hospital is defined as patients not needing to go to the hospital or clinic as often due to telehealth, and technology as a facilitator is defined as technology making healthcare feel more accessible for the patient. The most commonly cited subtheme is fewer trips to the hospital and was found in all 8 papers.

#### Privacy & Confidentiality

There were both negative and positive connotations to the theme privacy and confidentiality and they were coded as positive experiences (+) or negative experiences (-). Privacy and confidentiality as a positive experience (n=2) is defined as the feeling that private health information is protected when using telemedicine. A negative experience (n=3) is defined as the lack of privacy and confidentiality regarding health information.

#### Trust

Trust was organized into the subthemes comfort and relationship, with both having negative and positive subdivisions captured through coding. Positive views on comfort (n=3) include patients being comfortable receiving medical care from the team via telemedicine, whereas a negative view (n=3) of comfort might indicate patient discomfort. A positive view on relationship (n=3) indicated that patients felt telemedicine facilitated the patient-provider relationship, whereas negative views (n=4) suggested it hindered it.

## Discussion

The purpose of this study was to investigate the perspectives that kidney disease and transplant patients have regarding telemedicine. Through our qualitative meta-analysis we found that telemedicine has both strengths and limitations. As a strength, it is clear that the positive aspects of autonomy (positive engagement, motivation, and personalized) indicate that telemedicine provides an opportunity for patients to take an active part in their care. As was found in the paper by Huuskes et al., patients can sometimes feel overwhelmed by the hospital environment [18].

With distractions occurring in the hospital, it can be difficult for patients to focus on the health visit. Via telemedicine, patients can be at the comfort of their home and stay more engaged with their care team. Additionally, Kelly et al.’s group found that supportive text messages and phone calls encouraged patients to take more accountability for their diet [19]. The one negative attribute for autonomy (negative engagement) did however appear in the Trace et al. and Huuskes et al. papers, indicating that telemedicine did, at times, negatively affect patient experiences with engagement. In the Trace et al. paper, it was noted that pre-school children being involved in telemedicine understandably had a chaotic effect [20]. This suggests that while telemedicine can be promising, it is worth considering the age of the population that would best benefit from it. Notably, prior work has shown younger adult patients to be more satisfied with telemedicine when compared to older adults [21]. Therefore, the age of both pediatric and adult patients should be considered when deciding to implement telemedicine. Additionally, environment can affect engagement. In Huuskes et al.’s work, one patient indicated distractions in their environment when utilizing telemedicine, which negatively affected engagement [18]. Thus, before telemedicine is utilized, providers should discuss potential barriers, such as age and environment, with their patients.

This meta-analysis also elucidated that patients experienced both positive and negative logistical experiences with telemedicine. The most common logistical facilitator was fewer trips to the hospital, which was a theme found in every article included in the analysis. This indicates that one of telemedicine’s greatest strengths for patients with kidney disease or kidney transplant was tackling the issue of distance to and from the hospital or clinic. By having a virtual session at the comfort of home or work, patients could save a significant amount of time, which was appreciated by patients that lived both near to and far from the hospital [22]. This study found technology as both a barrier and a facilitator. As a barrier for example, in Neilson et al.’s work, patients indicated that virtual communication was challenging due to a lack of non-verbal cues such as “…body language and visual input” [23]. Comfort at home indicated that patients felt that telemedicine provided the opportunity to connect with the medical team in a more comfortable environment. Care as a logistical barrier suggested that telemedicine made patients feel concern over not having a physical exam [24]. They felt that by not being seen in person, the physician could not directly observe any issues that patients may present with physically such as edema [24]. Patients also felt that taking measurements at home such as blood pressure may not be as accurate as in the office [25].

This study demonstrated how patients perceived privacy in telemedicine positively and negatively. Positive views included patients feeling an elevated level of security [20]. This was because in clinic, patients felt that anyone could walk in and out of clinic, and that hallway conversations were not as secure. At home via telemedicine, however, patients felt the conversations to be more confidential. For negative views, patients felt that it could be challenging to locate a quiet area in the home or workplace for the appointment [22].

For positive views on comfort, Warner et al.’s work demonstrates that patients felt seen when they received personalized telehealth messages regarding their care [26]. They also felt comfortable discussing fears with the medical team via telephone consultation [26]. Warner et al.’s work also demonstrated positive views on relationship, with patients stating they were able to develop meaningful relationships with their coaches during the study [26]. However, in Neilson et al.’s work, it was noted that body language added nuance which could only really be done in person [23].

This current study shows how telemedicine contains both positive and negative attributes, as noted by the patients and caretakers it aims to serve. Subthemes and subdivisions that were cited in more of the 8 papers were deemed to be more meaningful characteristics. Based on the results of this study, it is our recommendation that telemedicine continue to be utilized and further studied. The benefits of this modality appear to make a great difference on patient access to care. Furthermore, the weaknesses of telemedicine should be addressed to provide the best experience for patients. This work demonstrates that telemedicine should be offered as an option to patients who are interested, and this modality is one hospital systems and clinics should consider implementing into practice.

There are some important nuances of note with regards to this study. Firstly, patient caretakers were considered a part of the study in interest. Given their critical role in caring for the patient, the researchers of this study believed that the telemedicine modalities meant to aid patient care directly applies to them as well. Secondly, studies that only discussed telemedicine modalities in a theoretical sense (e.g., patients making assumptions on telemedicine without relevant experience) were not utilized. While they are important to review to gauge interest in telemedicine, this study was geared more towards the effectiveness of telemedicine, rather than interest in using it. Finally, across the 8 studies selected, multiple modalities of telemedicine were used, including virtual visits, personalized messages, and telephone consultations. This is important to note given that different modalities have different strengths and weaknesses. For example, while telephone consultations open availability to conversations and saving time to drive to the hospital or clinic, they lack the visual component of a visit. Different software may also have different bugs that need addressing. The goal of this study was to focus on broad characteristics of telemedicine, rather than focusing on the nuanced minutia of each potential program.

The strengths of this study include a broad pediatric and adult population of both chronic kidney disease and transplant patients, multiple modalities of telemedicine, and high-quality works to assess telemedicine perspectives. Additionally, an enhancing transparency in reporting the synthesis of qualitative research (ENTREQ) checklist found originally in prior work was utilized to guide the methodology description in the writing of this manuscript (Supplemental Item 3) [27, 28]. Limitations of this study include a lack of provider perspective, the exclusion of non-English papers, and papers published before the year 2000. Additionally, since the start of this study, other papers have been published with regards to the field that should be considered as telemedicine continues to be utilized and potentially evolve. Papers that were published after data extraction began were also not included. Further studies should be performed to understand how the negative views of telemedicine can most effectively be addressed, and the most cost-effective way to implement secure telemedicine modalities into the healthcare setting.

## Conclusion

In conclusion, telemedicine offers a potential solution to help make medical appointments more accessible for chronic kidney disease and transplant patients. With telemedicine, there are both strengths and weaknesses as compared to in-person consultation. Of all characteristics of telemedicine studied here, avoiding travel to the hospital was the most cited view and was found in all 8 papers suggested. The authors recommend that telemedicine continue to be utilized and that negative characteristics of telemedicine continue to be studied for ways to improve the patient care experience. Future research should investigate ways to improve telemedicine use for patients such as improving perceived engagement, logistics, perceived confidentiality, and trust.

## Supporting information

Supplemental Item 1

Supplemental Item 2

Supplemental Item 3

## Data Availability

All data produced in the present study are available upon reasonable request to the authors.

## Acknowledgements

The authors would like to acknowledge Geisinger Commonwealth School of Medicine for its support of this research. Additionally, the authors would like to thank Iris Johnston and Amy Houck for their help with the literature search of this study.

## Online Supplemental Materials

**Supplemental Item 1**. The search terms for PubMed, Embase, and Academic Science Premier are listed

**Supplemental Item 2**. Crowe Critical Appraisal Tool for all 8 selected papers

**Supplemental Item 3**. ENTREQ Checklist

