## Supplemental Item 1 for "Telemedicine Perspectives of Patients with Non-Dialysis Kidney Disease and Kidney Transplant – A Qualitative Meta-Analysis"

**PubMed:**

renal replacement therapy OR kidney disease OR chronic kidney disease OR kidney failure OR chronic kidney failure OR mild renal impairment OR stage 1 kidney disease OR moderate renal impairment OR severe renal impairment OR end stage renal disease OR kidney transplantation OR hemodialysis OR haemodialysis OR hemofiltration OR haemofiltration OR hemodiafiltration OR haemodiafiltration OR dialysis OR Renal disease OR Renal failure OR Predialysis OR Pre-dialysis OR Kidney graft OR Renal graft OR Kidney allograft OR Renal allograft OR RRT OR Peritoneal dialysis OR HD

AND

Telehealth OR Telemedicine OR e-health OR ehealth OR m-health OR mhealth OR Remote consultation OR Remote care OR Video consultation OR Telephone consultation OR mobile device OR mobile application

AND

Attitudes OR Perspectives OR Experiences OR Perception OR Opinions OR Thoughts OR Feelings OR Beliefs OR viewpoints OR outlook

**EMBASE:**

Search Terms

(renal-replacement-therapy OR kidney-disease OR chronic-kidney-disease OR kidney-failure OR chronic-kidney-failure OR mild-renal-impairment OR stage-1-kidney-disease OR moderate-renal-impairment OR severe-renal-impairment OR end-stage-renal-disease OR kidney-transplantation OR hemodialysis OR haemodialysis OR hemofiltration OR haemofiltration OR hemodiafiltration OR haemodiafiltration OR dialysis OR Renal-disease OR Renal-failure OR Predialysis OR Pre-dialysis OR Kidney-graft OR Renal-graft OR Kidney-allograft OR Renal-allograft OR RRT OR Peritoneal-dialysis OR HD)

AND

(Telehealth OR Telemedicine OR e-health OR ehealth OR m-health OR mhealth OR Remote-consultation OR Remote-care OR Video-consultation OR Telephone-consultation OR mobile-device OR mobile-application)

AND

(Attitudes OR Perspectives OR Experiences OR Perception OR Opinions OR Thoughts OR Feelings OR Beliefs OR viewpoints OR outlook)

**Academic Search Premier:**

Search Terms:

renal replacement therapy OR kidney disease OR chronic kidney disease OR kidney failure OR chronic kidney failure OR mild renal impairment OR stage 1 kidney disease OR moderate renal impairment OR severe renal impairment OR end stage renal disease OR kidney transplantation OR hemodialysis OR haemodialysis OR hemofiltration OR haemofiltration OR hemodiafiltration OR haemodiafiltration OR dialysis OR Renal disease OR Renal failure OR Predialysis OR Pre-dialysis OR Kidney graft OR Renal graft OR Kidney allograft OR Renal allograft OR RRT OR Peritoneal dialysis OR HD

AND

Telehealth OR Telemedicine OR e-health OR ehealth OR m-health OR mhealth OR Remote consultation OR Remote care OR Video consultation OR Telephone consultation OR mobile device OR mobile application

AND

Attitudes OR Perspectives OR Experiences OR Perception OR Opinions OR Thoughts OR Feelings OR Beliefs OR viewpoints OR outlook
