## Supplemental Item 2 for "Telemedicine Perspectives of Patients with Non-Dialysis Kidney Disease and Kidney Transplant – A Qualitative Meta-Analysis"

#### Crowe Critical Appraisal Tool (CCAT) Form (v1.4)

Reference: https://doi.org/10.1177%2F20543581211066720 | Reviewer: CDM and ARC

This form must be used in conjunction with the CCAT User Guide (v1.4); otherwise validity and reliability may be severely compromised.

##### Citation

| Citation | Year |
|---|---|
| A Quantitative and Qualitative Study on Patient and Physician Perceptions of Nephrology Telephone Consultation During COVID-19 | 2022 |

##### Research design (add if not listed)

| | | |
|---|---|---|
| ❑ Not research | Article \| Editorial \| Report \| Opinion \| Guideline \| Pamphlet \| … | |
| ❑ Historical | … | |
| ☑ Qualitative | Narrative \| Phenomenology \| Ethnography \| Grounded theory \| Narrative case study \| … thematic analysis | |
| ☑ Descriptive, Exploratory, Observational | A. Cross-sectional \| Longitudinal \| Retrospective \| Prospective \| Correlational \| Predictive \| … | |
| | B. Cohort \| Case-control \| Survey \| Developmental \| Normative \| Case study \| … | |
| Experimental | ❑ True experiment | Pre-test/post-test control group \| Solomon four-group \| Post-test only control group \| Randomised two-factor \| Placebo controlled trial \| … |
| | ❑ Quasi-experiment | Post-test only \| Non-equivalent control group \| Counter balanced *(cross-over)* \| Multiple time series \| Separate sample pre-test post-test [no Control] [Control] \| … |
| | ❑ Single system | One-shot experimental *(case study)* \| Simple time series \| One group pre-test/post-test \| Interactive \| Multiple baseline \| Within subjects *(Equivalent time, repeated measures, multiple treatment)* \| … |
| ❑ Mixed Methods | Action research \| Sequential \| Concurrent \| Transformative \| … | |
| ❑ Synthesis | Systematic review \| Critical review \| Thematic synthesis \| Meta-ethnography \| Narrative synthesis \| … | |
| ❑ Other | … | |

##### Variables and analysis

| Intervention(s), Treatment(s), Exposure(s) | Outcome(s), Output(s), Predictor(s), Measure(s) | Data analysis method(s) |
|---|---|---|
| survey | patient and provider views on telephone consultations | descriptive statistics and content analysis |

##### Sampling

| Total size | | Group 1 | | Group 2 | | Group 3 | | Group 4 | | Control |
|---|---|---|---|---|---|---|---|---|---|---|
| Total size | 246 | Group 1 | 235 | Group 2 | 11 | Group 3 | | Group 4 | | Control |

Population, sample, setting: 235 patients and 11 physicians completed the survey

##### Data collection (add if not listed)

| | | | |
|---|---|---|---|
| Audit/Review | a) Primary \| Secondary \| …<br>b) Authoritative \| Partisan \| Antagonist \| …<br>c) Literature \| Systematic \| … | Interview | a) Formal \| Informal \| …<br>b) Structured \| Semi-structured \| Unstructured \| …<br>c) One-on-one \| Group \| Multiple \| Self-administered \| … |
| Observation | a) Participant \| Non-participant \| …<br>b) Structured \| Semi-structured \| Unstructured \| …<br>c) Covert \| Candid \| … | Testing | a) Standardised \| Norm-ref \| Criterion-ref \| Ipsative \| …<br>b) Objective \| Subjective \| …<br>c) One-on-one \| Group \| Self-administered \| … |

##### Scores

| | | | | | | | | | |
|---|---|---|---|---|---|---|---|---|---|
| Preliminaries | 5 | Design | 3 | Data Collection | 5 | Results | 5 | Total [/40] | 35 |
| Introduction | 5 | Sampling | 4 | Ethical Matters | 3 | Discussion | 5 | Total [%] | 87.5 |

##### General notes

CC BY-SA Crowe Critical Appraisal Tool (CCAT) :: Version 1.4 (19 November 2013) :: Michael Crowe
This work is licensed under the Creative Commons Attribution-ShareAlike 3.0 Unported License. To view a copy of this license, visit http://creativecommons.org/licenses/by-sa/3.0/

Appraise research on the merits of the research design used, not against other research designs.

| Category<br>Item | Item descriptors<br>[☑ Present; ☒ Absent; ■ Not applicable] | Description<br>[Important information for each item] | Score<br>[0–5] |
|---|---|---|---|
| **1. Preliminaries** | | | |
| Title | 1. Includes study aims ☑ and design ☑ | | |
| Abstract<br>(assess last) | 1. Key information ☑<br>2. Balanced ☑ and informative ☑ | | |
| Text<br>(assess last) | 1. Sufficient detail others could reproduce ☑<br>2. Clear/concise writing ☑, table(s) ☑, diagram(s) ■, figure(s) ☑ | | |
| | | **Preliminaries [/5]** | 5 |
| **2. Introduction** | | | |
| Background | 1. Summary of current knowledge ☑<br>2. Specific problem(s) addressed ☑ and reason(s) for addressing ☑ | | |
| Objective | 1. Primary objective(s), hypothesis(es), or aim(s) ☑<br>2. Secondary question(s) ■ | | |
| | **Is it worth continuing?** | **Introduction [/5]** | 5 |
| **3. Design** | | | |
| Research design | 1. Research design(s) chosen ☑ and why ☑<br>2. Suitability of research design(s) ☑ | | |
| Intervention, Treatment, Exposure | 1. Intervention(s)/treatment(s)/exposure(s) chosen ☑ and why ☒<br>2. Precise details of the intervention(s)/treatment(s)/exposure(s) ☑ for each group ☑<br>3. Intervention(s)/treatment(s)/exposure(s) valid ☒ and reliable ☒ | | |
| Outcome, Output, Predictor, Measure | 1. Outcome(s)/output(s)/predictor(s)/measure(s) chosen ☑ and why ☒<br>2. Clearly define outcome(s)/output(s)/predictor(s)/measure(s) ☑<br>3. Outcome(s)/output(s)/predictor(s)/measure(s) valid ☒ and reliable ☒ | | |
| Bias, etc | 1. Potential bias ☑, confounding variables ☒, effect modifiers ☒, interactions ☒<br>2. Sequence generation ■, group allocation ☑, group balance ☒, and by whom ☒<br>3. Equivalent treatment of participants/cases/groups ☒ | | |
| | **Is it worth continuing?** | **Design [/5]** | 3 |
| **4. Sampling** | | | |
| Sampling method | 1. Sampling method(s) chosen ☑ and why ☒<br>2. Suitability of sampling method ☒ | | |
| Sample size | 1. Sample size ☑, how chosen ☑, and why ☒<br>2. Suitability of sample size ☑ | | |
| Sampling protocol | 1. Target/actual/sample population(s): description ☑ and suitability ☑<br>2. Participants/cases/groups: inclusion ☑ and exclusion ☒ criteria<br>3. Recruitment of participants/cases/groups ☑ | | |
| | **Is it worth continuing?** | **Sampling [/5]** | 4 |
| **5. Data collection** | | | |
| Collection method | 1. Collection method(s) chosen ☑ and why ☒<br>2. Suitability of collection method(s) ☑ | | |
| Collection protocol | 1. Include date(s) ☑, location(s) ☑, setting(s) ☑, personnel ☑, materials ☑, processes ☑<br>2. Method(s) to ensure/enhance quality of measurement/instrumentation ☑<br>3. Manage non-participation ☒, withdrawal ☒, incomplete/lost data ☒ | | |
| | **Is it worth continuing?** | **Data collection [/5]** | 5 |
| **6. Ethical matters** | | | |
| Participant ethics | 1. Informed consent ☒, equity ☒<br>2. Privacy ☒, confidentiality/anonymity ☒ | only implied consent for nephrologists; mentioned follow-up study but didn't specify what this included | |
| Researcher ethics | 1. Ethical approval ☑, funding ☑, conflict(s) of interest ☑<br>2. Subjectivities ☒, relationship(s) with participants/cases ☒ | | |
| | **Is it worth continuing?** | **Ethical matters [/5]** | 3 |
| **7. Results** | | | |
| Analysis, Integration, Interpretation method | 1. A.I.I. method(s) for primary outcome(s)/output(s)/predictor(s) chosen ☑ and why ☒<br>2. Additional A.I.I. methods (e.g. subgroup analysis) chosen ■ and why ■<br>3. Suitability of analysis/integration/interpretation method(s) ☑ | | |
| Essential analysis | 1. Flow of participants/cases/groups through each stage of research ☑<br>2. Demographic and other characteristics of participants/cases/groups ☑<br>3. Analyse raw data ☑, response rate ☑, non-participation/withdrawal/incomplete/lost data ☑ | | |
| Outcome, Output, Predictor analysis | 1. Summary of results ☑ and precision ☒ for each outcome/output/predictor/measure<br>2. Consideration of benefits/harms ☑, unexpected results ■, problems/failures ■<br>3. Description of outlying data (e.g. diverse cases, adverse effects, minor themes) ■ | data reported in percentages, but does not explain differences in completion of survey questions | |
| | | **Results [/5]** | 5 |
| **8. Discussion** | | | |
| Interpretation | 1. Interpretation of results in the context of current evidence ☑ and objectives ☑<br>2. Draw inferences consistent with the strength of the data ☑<br>3. Consideration of alternative explanations for observed results ☑<br>4. Account for bias ☑, confounding/effect modifiers/interactions/imprecision ☑ | | |
| Generalisation | 1. Consideration of overall practical usefulness of the study ☑<br>2. Description of generalisability (external validity) of the study ☑ | | |
| Concluding remarks | 1. Highlight study's particular strengths ☒<br>2. Suggest steps that may improve future results (e.g. limitations) ☑<br>3. Suggest further studies ☑ | | |
| | | **Discussion [/5]** | 5 |
| **9. Total** | | | |
| Total score | 1. Add all scores for categories 1–8 | | |
| | | **Total [/40]** | 35 |

#### Crowe Critical Appraisal Tool (CCAT) Form (v1.4)

Reference: doi: 10.1111/tri.13934 Reviewer: CDM and ARC

This form must be used in conjunction with the CCAT User Guide (v1.4); otherwise validity and reliability may be severely compromised.

##### Citation

| Citation | Year |
|---|---|
| Kidney transplant recipient perspectives on telehealth during the COVID-19 pandemic | 2021 |

##### Research design (add if not listed)

| | | |
|---|---|---|
| ❑ Not research | | Article \| Editorial \| Report \| Opinion \| Guideline \| Pamphlet \| … |
| ❑ Historical | | … |
| ☑ Qualitative | | Narrative \| Phenomenology \| Ethnography \| Grounded theory \| Narrative case study \| … thematic analysis |
| ❑ Descriptive, Exploratory, Observational | | A. Cross-sectional \| Longitudinal \| Retrospective \| Prospective \| Correlational \| Predictive \| … |
| | | B. Cohort \| Case-control \| Survey \| Developmental \| Normative \| Case study \| … |
| Experimental | ❑ True experiment | Pre-test/post-test control group \| Solomon four-group \| Post-test only control group \| Randomised two-factor \| Placebo controlled trial \| … |
| | ❑ Quasi-experiment | Post-test only \| Non-equivalent control group \| Counter balanced *(cross-over)* \| Multiple time series \| Separate sample pre-test post-test [no Control] [Control] \| … |
| | ❑ Single system | One-shot experimental *(case study)* \| Simple time series \| One group pre-test/post-test \| Interactive \| Multiple baseline \| Within subjects *(Equivalent time, repeated measures, multiple treatment)* \| … |
| ❑ Mixed Methods | | Action research \| Sequential \| Concurrent \| Transformative \| … |
| ❑ Synthesis | | Systematic review \| Critical review \| Thematic synthesis \| Meta-ethnography \| Narrative synthesis \| … |
| ❑ Other | | … |

##### Variables and analysis

| Intervention(s), Treatment(s), Exposure(s) | Outcome(s), Output(s), Predictor(s), Measure(s) | Data analysis method(s) |
|---|---|---|
| focus groups | kidney transplant perspectives | thematic analysis |

##### Sampling

| Total size | Group 1 | Group 2 | Group 3 | Group 4 | Control |
|---|---|---|---|---|---|
| 34 | | | | | |

Population, sample, setting:

##### Data collection (add if not listed)

| | | | |
|---|---|---|---|
| Audit/Review | a) Primary \| Secondary \| …<br>b) Authoritative \| Partisan \| Antagonist \| …<br>c) Literature \| Systematic \| … | Interview | a) Formal \| Informal \| …<br>b) Structured \| Semi-structured \| Unstructured \| …<br>c) One-on-one \| Group \| Multiple \| Self-administered \| … |
| Observation | a) Participant \| Non-participant \| …<br>b) Structured \| Semi-structured \| Unstructured \| …<br>c) Covert \| Candid \| … | Testing | a) Standardised \| Norm-ref \| Criterion-ref \| Ipsative \| …<br>b) Objective \| Subjective \| …<br>c) One-on-one \| Group \| Self-administered \| … |

##### Scores

| | | | | | | | | | |
|---|---|---|---|---|---|---|---|---|---|
| Preliminaries | 5 | Design | 4 | Data Collection | 4 | Results | 5 | Total [/40] | 38 |
| Introduction | 5 | Sampling | 5 | Ethical Matters | 5 | Discussion | 5 | Total [%] | 95 |

##### General notes

Crowe Critical Appraisal Tool (CCAT) :: Version 1.4 (19 November 2013) :: Michael Crowe
This work is licensed under the Creative Commons Attribution-ShareAlike 3.0 Unported License. To view a copy of this license, visit http://creativecommons.org/licenses/by-sa/3.0/

Appraise research on the merits of the research design used, not against other research designs.

| Category<br>Item | Item descriptors<br>[☑ Present; ☒ Absent; ■ Not applicable] | Description<br>[Important information for each item] | Score<br>[0–5] |
|---|---|---|---|
| **1. Preliminaries** | | | |
| Title | 1. Includes study aims ☑ and design ☒ | | |
| Abstract<br>(assess last) | 1. Key information ☑<br>2. Balanced ☑ and informative ☑ | | |
| Text<br>(assess last) | 1. Sufficient detail others could reproduce ☑<br>2. Clear/concise writing ☑, table(s) ☑, diagram(s) ■, figure(s) ☑ | | |
| | | **Preliminaries [/5]** | 5 |
| **2. Introduction** | | | |
| Background | 1. Summary of current knowledge ☑<br>2. Specific problem(s) addressed ☑ and reason(s) for addressing ☑ | | |
| Objective | 1. Primary objective(s), hypothesis(es), or aim(s) ☑<br>2. Secondary question(s) ■ | | |
| | **Is it worth continuing?** | **Introduction [/5]** | 5 |
| **3. Design** | | | |
| Research design | 1. Research design(s) chosen ☑ and why ☑<br>2. Suitability of research design(s) ☑ | | |
| Intervention, Treatment, Exposure | 1. Intervention(s)/treatment(s)/exposure(s) chosen ☑ and why ☑<br>2. Precise details of the intervention(s)/treatment(s)/exposure(s) ☑ for each group ■<br>3. Intervention(s)/treatment(s)/exposure(s) valid ☒ and reliable ☒ | | |
| Outcome, Output, Predictor, Measure | 1. Outcome(s)/output(s)/predictor(s)/measure(s) chosen ☑ and why ☒<br>2. Clearly define outcome(s)/output(s)/predictor(s)/measure(s) ☑<br>3. Outcome(s)/output(s)/predictor(s)/measure(s) valid ☒ and reliable ☒ | | |
| Bias, etc | 1. Potential bias ☑, confounding variables ☒, effect modifiers ☒, interactions ☒<br>2. Sequence generation ■, group allocation ■, group balance ■, and by whom ■<br>3. Equivalent treatment of participants/cases/groups ☑ | | |
| | **Is it worth continuing?** | **Design [/5]** | 4 |
| **4. Sampling** | | | |
| Sampling method | 1. Sampling method(s) chosen ☑ and why ☒<br>2. Suitability of sampling method ☑ | | |
| Sample size | 1. Sample size ☑, how chosen ☑, and why ☑<br>2. Suitability of sample size ☑ | | |
| Sampling protocol | 1. Target/actual/sample population(s): description ☑ and suitability ☑<br>2. Participants/cases/groups: inclusion ☑ and exclusion ☑ criteria<br>3. Recruitment of participants/cases/groups ☑ | | |
| | **Is it worth continuing?** | **Sampling [/5]** | 5 |
| **5. Data collection** | | | |
| Collection method | 1. Collection method(s) chosen ☑ and why ☒<br>2. Suitability of collection method(s) ☑ | | |
| Collection protocol | 1. Include date(s) ☑, location(s) ☑, setting(s) ☑, personnel ☑, materials ☑, processes ☑<br>2. Method(s) to ensure/enhance quality of measurement/instrumentation ☑<br>3. Manage non-participation ☒, withdrawal ☒, incomplete/lost data ☒ | | |
| | **Is it worth continuing?** | **Data collection [/5]** | 4 |
| **6. Ethical matters** | | | |
| Participant ethics | 1. Informed consent ☑, equity ☒<br>2. Privacy ■, confidentiality/anonymity ■ | | |
| Researcher ethics | 1. Ethical approval ☑, funding ☑, conflict(s) of interest ☑<br>2. Subjectivities ☒, relationship(s) with participants/cases ☒ | | |
| | **Is it worth continuing?** | **Ethical matters [/5]** | 5 |
| **7. Results** | | | |
| Analysis, Integration, Interpretation method | 1. A.I.I. method(s) for primary outcome(s)/output(s)/predictor(s) chosen ☑ and why ☒<br>2. Additional A.I.I. methods (e.g. subgroup analysis) chosen ■ and why ■<br>3. Suitability of analysis/integration/interpretation method(s) ☑ | | |
| Essential analysis | 1. Flow of participants/cases/groups through each stage of research ☑<br>2. Demographic and other characteristics of participants/cases/groups ☑<br>3. Analyse raw data ☑, response rate ☑, non-participation/withdrawal/incomplete/lost data ☒ | | |
| Outcome, Output, Predictor analysis | 1. Summary of results ☑ and precision ■ for each outcome/output/predictor/measure<br>2. Consideration of benefits/harms ☑, unexpected results ■, problems/failures ■<br>3. Description of outlying data (e.g. diverse cases, adverse effects, minor themes) ☑ | | |
| | | **Results [/5]** | 5 |
| **8. Discussion** | | | |
| Interpretation | 1. Interpretation of results in the context of current evidence ☑ and objectives ☑<br>2. Draw inferences consistent with the strength of the data ☑<br>3. Consideration of alternative explanations for observed results ■<br>4. Account for bias ☑, confounding/effect modifiers/interactions/imprecision ☑ | | |
| Generalisation | 1. Consideration of overall practical usefulness of the study ☑<br>2. Description of generalisability (external validity) of the study ☑ | | |
| Concluding remarks | 1. Highlight study's particular strengths ☑<br>2. Suggest steps that may improve future results (e.g. limitations) ☑<br>3. Suggest further studies ☑ | | |
| | | **Discussion [/5]** | 5 |
| **9. Total** | | | |
| Total score | 1. Add all scores for categories 1–8 | | |
| | | **Total [/40]** | 38 |

### Crowe Critical Appraisal Tool (CCAT) Form (v1.4)

Reference: doi:10.1136/ bmjopen-2018-024551

Reviewer: CDM and ARC

This form must be used in conjunction with the CCAT User Guide (v1.4); otherwise validity and reliability may be severely compromised.

#### Citation

| Citation | Year |
|---|---|
| Feasibility and acceptability of telehealth coaching to promote healthy eating in chronic kidney disease: a mixed-methods process evaluation | 2019 |

#### Research design (add if not listed)

| | | |
|---|---|---|
| ❑ Not research | | Article \| Editorial \| Report \| Opinion \| Guideline \| Pamphlet \| … |
| ❑ Historical | | … |
| ❑ Qualitative | | Narrative \| Phenomenology \| Ethnography \| Grounded theory \| Narrative case study \| … |
| ❑ Descriptive, Exploratory, Observational | | A. Cross-sectional \| Longitudinal \| Retrospective \| Prospective \| Correlational \| Predictive \| … |
| | | B. Cohort \| Case-control \| Survey \| Developmental \| Normative \| Case study \| … |
| Experimental | ❑ True experiment | Pre-test/post-test control group \| Solomon four-group \| Post-test only control group \| Randomised two-factor \| Placebo controlled trial \| … |
| | ❑ Quasi-experiment | Post-test only \| Non-equivalent control group \| Counter balanced *(cross-over)* \| Multiple time series \| Separate sample pre-test post-test [no Control] [Control] \| … |
| | ❑ Single system | One-shot experimental *(case study)* \| Simple time series \| One group pre-test/post-test \| Interactive \| Multiple baseline \| Within subjects *(Equivalent time, repeated measures, multiple treatment)* \| … |
| ☑ Mixed Methods | | Action research \| Sequential \| Concurrent \| Transformative \| … mixed-methods process evaluation embedded in a RCT |
| ❑ Synthesis | | Systematic review \| Critical review \| Thematic synthesis \| Meta-ethnography \| Narrative synthesis \| … |
| ❑ Other | | … |

#### Variables and analysis

| Intervention(s), Treatment(s), Exposure(s) | Outcome(s), Output(s), Predictor(s), Measure(s) | Data analysis method(s) |
|---|---|---|
| telephone calls and personalized texts | feasibility and acceptability of telehealth model | descriptive statistics and qualitative analysis |

#### Sampling

| Total size | Group 1 | Group 2 | Group 3 | Group 4 | Control |
|---|---|---|---|---|---|
| 80 | 41 | 39 | | | |

Population, sample, setting: patients with chronic kidney disease stages 3-4
group 1 = experimental group 2 = control

#### Data collection (add if not listed)

| | | | |
|---|---|---|---|
| Audit/Review | a) Primary \| Secondary \| …<br>b) Authoritative \| Partisan \| Antagonist \| …<br>c) Literature \| Systematic \| … | Interview | a) Formal \| Informal \| …<br>b) Structured \| Semi-structured \| Unstructured \| …<br>c) One-on-one \| Group \| Multiple \| Self-administered \| … |
| Observation | a) Participant \| Non-participant \| …<br>b) Structured \| Semi-structured \| Unstructured \| …<br>c) Covert \| Candid \| … | Testing | a) Standardised \| Norm-ref \| Criterion-ref \| Ipsative \| …<br>b) Objective \| Subjective \| …<br>c) One-on-one \| Group \| Self-administered \| … |

#### Scores

| | | | | | | | | | |
|---|---|---|---|---|---|---|---|---|---|
| Preliminaries | 5 | Design | 4 | Data Collection | 5 | Results | 5 | Total [/40] | 38 |
| Introduction | 5 | Sampling | 5 | Ethical Matters | 4 | Discussion | 5 | Total [%] | 95 |

#### General notes

BY-SA Crowe Critical Appraisal Tool (CCAT) :: Version 1.4 (19 November 2013) :: Michael Crowe
This work is licensed under the Creative Commons Attribution-ShareAlike 3.0 Unported License. To view a copy of this license, visit http://creativecommons.org/licenses/by-sa/3.0/

Appraise research on the merits of the research design used, not against other research designs.

| Category<br>Item | Item descriptors<br>[☑ Present; ☒ Absent; ■ Not applicable] | Description<br>[Important information for each item] | Score<br>[0–5] |
|---|---|---|---|
| **1. Preliminaries** | | | |
| Title | 1. Includes study aims ☑ and design ☑ | | |
| Abstract<br>(assess last) | 1. Key information ☑<br>2. Balanced ☑ and informative ☑ | | |
| Text<br>(assess last) | 1. Sufficient detail others could reproduce ❑<br>2. Clear/concise writing ☑, table(s) ☑, diagram(s) ■, figure(s) ☑ | | |
| | | **Preliminaries [/5]** | 5 |
| **2. Introduction** | | | |
| Background | 1. Summary of current knowledge ☑<br>2. Specific problem(s) addressed ☑ and reason(s) for addressing ☑ | | |
| Objective | 1. Primary objective(s), hypothesis(es), or aim(s) ☑<br>2. Secondary question(s) ■ | | |
| | **Is it worth continuing?** | **Introduction [/5]** | 5 |
| **3. Design** | | | |
| Research design | 1. Research design(s) chosen ☑ and why ☑<br>2. Suitability of research design(s) ☑ | | |
| Intervention, Treatment, Exposure | 1. Intervention(s)/treatment(s)/exposure(s) chosen ☑ and why ☒<br>2. Precise details of the intervention(s)/treatment(s)/exposure(s) ☑ for each group ☑<br>3. Intervention(s)/treatment(s)/exposure(s) valid ☑ and reliable ☑ | | |
| Outcome, Output, Predictor, Measure | 1. Outcome(s)/output(s)/predictor(s)/measure(s) chosen ☑ and why ☒<br>2. Clearly define outcome(s)/output(s)/predictor(s)/measure(s) ☑<br>3. Outcome(s)/output(s)/predictor(s)/measure(s) valid ☑ and reliable ☑ | | |
| Bias, etc | 1. Potential bias ☑, confounding variables ☒, effect modifiers ☒, interactions ☑<br>2. Sequence generation ☑, group allocation ☑, group balance ☑, and by whom ☑<br>3. Equivalent treatment of participants/cases/groups ☒ | | |
| | **Is it worth continuing?** | **Design [/5]** | 4 |
| **4. Sampling** | | | |
| Sampling method | 1. Sampling method(s) chosen ☑ and why ☒<br>2. Suitability of sampling method ☑ | | |
| Sample size | 1. Sample size ☑, how chosen ☑, and why ☑<br>2. Suitability of sample size ☑ | | |
| Sampling protocol | 1. Target/actual/sample population(s): description ☑ and suitability ☑<br>2. Participants/cases/groups: inclusion ☑ and exclusion ☑ criteria<br>3. Recruitment of participants/cases/groups ☑ | | |
| | **Is it worth continuing?** | **Sampling [/5]** | 5 |
| **5. Data collection** | | | |
| Collection method | 1. Collection method(s) chosen ☑ and why ☒<br>2. Suitability of collection method(s) ☑ | | |
| Collection protocol | 1. Include date(s) ☑, location(s) ☑, setting(s) ☑, personnel ☑, materials ☑, processes ☑<br>2. Method(s) to ensure/enhance quality of measurement/instrumentation ☑<br>3. Manage non-participation ☑, withdrawal ☑, incomplete/lost data ☑ | | |
| | **Is it worth continuing?** | **Data collection [/5]** | 5 |
| **6. Ethical matters** | | | |
| Participant ethics | 1. Informed consent ☑, equity ☑<br>2. Privacy ☒, confidentiality/anonymity ☒ | | |
| Researcher ethics | 1. Ethical approval ☑, funding ☑, conflict(s) of interest ☑<br>2. Subjectivities ☑, relationship(s) with participants/cases ☑ | | |
| | **Is it worth continuing?** | **Ethical matters [/5]** | 4 |
| **7. Results** | | | |
| Analysis, Integration, Interpretation method | 1. A.I.I. method(s) for primary outcome(s)/output(s)/predictor(s) chosen ☑ and why ☑<br>2. Additional A.I.I. methods (e.g. subgroup analysis) chosen ■ and why ■<br>3. Suitability of analysis/integration/interpretation method(s) ☑ | | |
| Essential analysis | 1. Flow of participants/cases/groups through each stage of research ☑<br>2. Demographic and other characteristics of participants/cases/groups ☑<br>3. Analyse raw data ☑, response rate ☑, non-participation/withdrawal/incomplete/lost data ☑ | | |
| Outcome, Output, Predictor analysis | 1. Summary of results ☑ and precision ☑ for each outcome/output/predictor/measure<br>2. Consideration of benefits/harms ☑, unexpected results ☑, problems/failures ☑<br>3. Description of outlying data (e.g. diverse cases, adverse effects, minor themes) ☑ | | |
| | | **Results [/5]** | 5 |
| **8. Discussion** | | | |
| Interpretation | 1. Interpretation of results in the context of current evidence ☑ and objectives ☑<br>2. Draw inferences consistent with the strength of the data ☑<br>3. Consideration of alternative explanations for observed results ☑<br>4. Account for bias ☒, confounding/effect modifiers/interactions/imprecision ☑ | | |
| Generalisation | 1. Consideration of overall practical usefulness of the study ☑<br>2. Description of generalisability (external validity) of the study ☑ | | |
| Concluding remarks | 1. Highlight study's particular strengths ☑<br>2. Suggest steps that may improve future results (e.g. limitations) ☑<br>3. Suggest further studies ☑ | | |
| | | **Discussion [/5]** | 5 |
| **9. Total** | | | |
| Total score | 1. Add all scores for categories 1–8 | | |
| | | **Total [/40]** | 38 |

### Crowe Critical Appraisal Tool (CCAT) Form (v1.4)

Reference: doi:10.1001/jamanetworkopen.2021.37193 | Reviewer: CDM and ARC

This form must be used in conjunction with the CCAT User Guide (v1.4); otherwise validity and reliability may be severely compromised.

#### Citation

| Citation | Year |
|---|---|
| Perceptions of Telehealth vs In-Person Visits Among Older Adults With Advanced Kidney Disease, Care Partners, and Clinicians | 2021 |

#### Research design (add if not listed)

| | | |
|---|---|---|
| ❑ Not research | | Article \| Editorial \| Report \| Opinion \| Guideline \| Pamphlet \| … |
| ❑ Historical | | … |
| ☑ Qualitative | | Narrative \| Phenomenology \| Ethnography \| Grounded theory \| Narrative case study \| … thematic analysis |
| ❑ Descriptive, Exploratory, Observational | | A. Cross-sectional \| Longitudinal \| Retrospective \| Prospective \| Correlational \| Predictive \| … |
| | | B. Cohort \| Case-control \| Survey \| Developmental \| Normative \| Case study \| … |
| Experimental | ❑ True experiment | Pre-test/post-test control group \| Solomon four-group \| Post-test only control group \| Randomised two-factor \| Placebo controlled trial \| … |
| | ❑ Quasi-experiment | Post-test only \| Non-equivalent control group \| Counter balanced *(cross-over)* \| Multiple time series \| Separate sample pre-test post-test [no Control] [Control] \| … |
| | ❑ Single system | One-shot experimental *(case study)* \| Simple time series \| One group pre-test/post-test \| Interactive \| Multiple baseline \| Within subjects *(Equivalent time, repeated measures, multiple treatment)* \| … |
| ❑ Mixed Methods | | Action research \| Sequential \| Concurrent \| Transformative \| … |
| ❑ Synthesis | | Systematic review \| Critical review \| Thematic synthesis \| Meta-ethnography \| Narrative synthesis \| … |
| ❑ Other | | … |

#### Variables and analysis

| Intervention(s), Treatment(s), Exposure(s) | Outcome(s), Output(s), Predictor(s), Measure(s) | Data analysis method(s) |
|---|---|---|
| telehealth | semi-structured interview | verbatim transcription and thematic analysis |

#### Sampling

| Total size | | Group 1 | | Group 2 | | Group 3 | | Group 4 | | Control |
|---|---|---|---|---|---|---|---|---|---|---|
| | 60 | | 19 | | 30 | | 11 | | | |

Population, sample, setting: group 1 - clinicians; group 2 - patients; group 3 - care partners

#### Data collection (add if not listed)

| | | | |
|---|---|---|---|
| Audit/Review | a) Primary \| Secondary \| …<br>b) Authoritative \| Partisan \| Antagonist \| …<br>c) Literature \| Systematic \| … | Interview | a) Formal \| Informal \| …<br>b) Structured \| Semi-structured \| Unstructured \| …<br>c) One-on-one \| Group \| Multiple \| Self-administered \| … |
| Observation | a) Participant \| Non-participant \| …<br>b) Structured \| Semi-structured \| Unstructured \| …<br>c) Covert \| Candid \| … | Testing | a) Standardised \| Norm-ref \| Criterion-ref \| Ipsative \| …<br>b) Objective \| Subjective \| …<br>c) One-on-one \| Group \| Self-administered \| … |

#### Scores

| | | | | | | | | | |
|---|---|---|---|---|---|---|---|---|---|
| Preliminaries | 4 | Design | 4 | Data Collection | 4 | Results | 5 | Total [/40] | 36 |
| Introduction | 5 | Sampling | 5 | Ethical Matters | 4 | Discussion | 5 | Total [%] | 90 |

#### General notes

CC BY-SA Crowe Critical Appraisal Tool (CCAT) :: Version 1.4 (19 November 2013) :: Michael Crowe
This work is licensed under the Creative Commons Attribution-ShareAlike 3.0 Unported License. To view a copy of this license, visit http://creativecommons.org/licenses/by-sa/3.0/

Appraise research on the merits of the research design used, not against other research designs.

| Category<br>Item | Item descriptors<br>[☑ Present; ☒ Absent; ■ Not applicable] | Description<br>[Important information for each item] | Score<br>[0–5] |
|---|---|---|---|
| **1. Preliminaries** | | | |
| Title | 1. Includes study aims ☑ and design ☒ | | |
| Abstract<br>(assess last) | 1. Key information ☑<br>2. Balanced ☑ and informative ☑ | | |
| Text<br>(assess last) | 1. Sufficient detail others could reproduce ☑<br>2. Clear/concise writing ☑, table(s) ☑, diagram(s) ☒, figure(s) ☒ | | |
| | | **Preliminaries [/5]** | 4 |
| **2. Introduction** | | | |
| Background | 1. Summary of current knowledge ☑<br>2. Specific problem(s) addressed ☑ and reason(s) for addressing ☑ | | |
| Objective | 1. Primary objective(s), hypothesis(es), or aim(s) ☑<br>2. Secondary question(s) ■ | | |
| | **Is it worth continuing?** | **Introduction [/5]** | 5 |
| **3. Design** | | | |
| Research design | 1. Research design(s) chosen ☑ and why ☑<br>2. Suitability of research design(s) ☑ | | |
| Intervention, Treatment, Exposure | 1. Intervention(s)/treatment(s)/exposure(s) chosen ☑ and why ☑<br>2. Precise details of the intervention(s)/treatment(s)/exposure(s) ☑ for each group ☑<br>3. Intervention(s)/treatment(s)/exposure(s) valid ☑ and reliable ☑ | | |
| Outcome, Output, Predictor, Measure | 1. Outcome(s)/output(s)/predictor(s)/measure(s) chosen ☑ and why ☑<br>2. Clearly define outcome(s)/output(s)/predictor(s)/measure(s) ☑<br>3. Outcome(s)/output(s)/predictor(s)/measure(s) valid ☑ and reliable ☑ | | |
| Bias, etc | 1. Potential bias ☑, confounding variables ■, effect modifiers ☑, interactions ☒<br>2. Sequence generation ■, group allocation ☑, group balance ■, and by whom ■<br>3. Equivalent treatment of participants/cases/groups ☑ | | |
| | **Is it worth continuing?** | **Design [/5]** | 4 |
| **4. Sampling** | | | |
| Sampling method | 1. Sampling method(s) chosen ☑ and why ☑<br>2. Suitability of sampling method ☑ | | |
| Sample size | 1. Sample size ☑, how chosen ☑, and why ☑<br>2. Suitability of sample size ☑ | | |
| Sampling protocol | 1. Target/actual/sample population(s): description ☑ and suitability ☑<br>2. Participants/cases/groups: inclusion ☑ and exclusion ☑ criteria<br>3. Recruitment of participants/cases/groups ☑ | | |
| | **Is it worth continuing?** | **Sampling [/5]** | 5 |
| **5. Data collection** | | | |
| Collection method | 1. Collection method(s) chosen ☑ and why ☑<br>2. Suitability of collection method(s) ☑ | | |
| Collection protocol | 1. Include date(s) ☑, location(s) ☑, setting(s) ☑, personnel ☑, materials ☑, processes ☑<br>2. Method(s) to ensure/enhance quality of measurement/instrumentation ☑<br>3. Manage non-participation ☒, withdrawal ☒, incomplete/lost data ☒ | | |
| | **Is it worth continuing?** | **Data collection [/5]** | 4 |
| **6. Ethical matters** | | | |
| Participant ethics | 1. Informed consent ☑, equity ☑<br>2. Privacy ☒, confidentiality/anonymity ☒ | each group has their own interview | |
| Researcher ethics | 1. Ethical approval ☑, funding ☑, conflict(s) of interest ☑<br>2. Subjectivities ☑, relationship(s) with participants/cases ☒ | uncertain what relationship between researchers and clinicians is | |
| | **Is it worth continuing?** | **Ethical matters [/5]** | 4 |
| **7. Results** | | | |
| Analysis, Integration, Interpretation method | 1. A.I.I. method(s) for primary outcome(s)/output(s)/predictor(s) chosen ☑ and why ☑<br>2. Additional A.I.I. methods (e.g. subgroup analysis) chosen ☑ and why ☑<br>3. Suitability of analysis/integration/interpretation method(s) ☑ | additional analysis done for differences and similarities of perspectives in race and ethnicity | |
| Essential analysis | 1. Flow of participants/cases/groups through each stage of research ☑<br>2. Demographic and other characteristics of participants/cases/groups ☑<br>3. Analyse raw data ☑, response rate ☒, non-participation/withdrawal/incomplete/lost data ☒ | | |
| Outcome, Output, Predictor analysis | 1. Summary of results ☑ and precision ☑ for each outcome/output/predictor/measure<br>2. Consideration of benefits/harms ☑, unexpected results ■, problems/failures ■<br>3. Description of outlying data (e.g. diverse cases, adverse effects, minor themes) ☑ | | |
| | | **Results [/5]** | 5 |
| **8. Discussion** | | | |
| Interpretation | 1. Interpretation of results in the context of current evidence ☑ and objectives ☑<br>2. Draw inferences consistent with the strength of the data ☑<br>3. Consideration of alternative explanations for observed results ■<br>4. Account for bias ☑, confounding/effect modifiers/interactions/imprecision ☑ | | |
| Generalisation | 1. Consideration of overall practical usefulness of the study ☑<br>2. Description of generalisability (external validity) of the study ☑ | | |
| Concluding remarks | 1. Highlight study's particular strengths ☑<br>2. Suggest steps that may improve future results (e.g. limitations) ☑<br>3. Suggest further studies ☑ | | |
| | | **Discussion [/5]** | 5 |
| **9. Total** | | | |
| Total score | 1. Add all scores for categories 1–8 | | |
| | | **Total [/40]** | 36 |

### Crowe Critical Appraisal Tool (CCAT) Form (v1.4)

Reference: DOI: 10.1111/jocn.15178 Reviewer: CDM and ARC

This form must be used in conjunction with the CCAT User Guide (v1.4); otherwise validity and reliability may be severely compromised.

#### Citation

| Citation | Year |
|---|---|
| Evaluation of a telehealth solution developed to improve follow-up after kidney Transplantation | 2019 |

#### Research design (add if not listed)

| | | |
|---|---|---|
| ❑ Not research | | Article \| Editorial \| Report \| Opinion \| Guideline \| Pamphlet \| ... |
| ❑ Historical | | ... |
| ☑ Qualitative | | Narrative \| Phenomenology \| Ethnography \| Grounded theory \| Narrative case study \| ... thematic analysis |
| ❑ Descriptive, Exploratory, Observational | | A. Cross-sectional \| Longitudinal \| Retrospective \| Prospective \| Correlational \| Predictive \| ... |
| | | B. Cohort \| Case-control \| Survey \| Developmental \| Normative \| Case study \| ... |
| Experimental | ❑ True experiment | Pre-test/post-test control group \| Solomon four-group \| Post-test only control group \| Randomised two-factor \| Placebo controlled trial \| ... |
| | ❑ Quasi-experiment | Post-test only \| Non-equivalent control group \| Counter balanced *(cross-over)* \| Multiple time series \| Separate sample pre-test post-test [no Control] [Control] \| ... |
| | ❑ Single system | One-shot experimental *(case study)* \| Simple time series \| One group pre-test/post-test \| Interactive \| Multiple baseline \| Within subjects *(Equivalent time, repeated measures, multiple treatment)* \| ... |
| ❑ Mixed Methods | | Action research \| Sequential \| Concurrent \| Transformative \| ... |
| ❑ Synthesis | | Systematic review \| Critical review \| Thematic synthesis \| Meta-ethnography \| Narrative synthesis \| ... |
| ❑ Other | | ... |

#### Variables and analysis

| Intervention(s), Treatment(s), Exposure(s) | Outcome(s), Output(s), Predictor(s), Measure(s) | Data analysis method(s) |
|---|---|---|
| telehealth intervention | patient and provider perspectives | semi-structured interviews |

#### Sampling

| | | | | | | | | | | |
|---|---|---|---|---|---|---|---|---|---|---|
| Total size | 36 | Group 1 | 16 | Group 2 | 20 | Group 3 | | Group 4 | | Control |
| Population, sample, setting | 16 patients and 20 healthcare professionas | | | | | | | | | |

#### Data collection (add if not listed)

| | | | |
|---|---|---|---|
| Audit/Review | a) Primary \| Secondary \| ...<br>b) Authoritative \| Partisan \| Antagonist \| ...<br>c) Literature \| Systematic \| ... | Interview | a) Formal \| Informal \| ...<br>b) Structured \| Semi-structured \| Unstructured \| ...<br>c) One-on-one \| Group \| Multiple \| Self-administered \| ... |
| Observation | a) Participant \| Non-participant \| ...<br>b) Structured \| Semi-structured \| Unstructured \| ...<br>c) Covert \| Candid \| ... | Testing | a) Standardised \| Norm-ref \| Criterion-ref \| Ipsative \| ...<br>b) Objective \| Subjective \| ...<br>c) One-on-one \| Group \| Self-administered \| ... |

#### Scores

| | | | | | | | | | |
|---|---|---|---|---|---|---|---|---|---|
| Preliminaries | 4 | Design | 4 | Data Collection | 5 | Results | 5 | Total [/40] | 34 |
| Introduction | 5 | Sampling | 4 | Ethical Matters | 3 | Discussion | 4 | Total [%] | 85 |

#### General notes

(cc) BY-SA Crowe Critical Appraisal Tool (CCAT) :: Version 1.4 (19 November 2013) :: Michael Crowe
This work is licensed under the Creative Commons Attribution-ShareAlike 3.0 Unported License. To view a copy of this license, visit http://creativecommons.org/licenses/by-sa/3.0/

Appraise research on the merits of the research design used, not against other research designs.

| Category<br>Item | Item descriptors<br>[☑ Present; ☒ Absent; ■ Not applicable] | Description<br>[Important information for each item] | Score<br>[0–5] |
|---|---|---|---|
| **1. Preliminaries** | | | |
| Title | 1. Includes study aims ☑ and design ☒ | | |
| Abstract<br>(assess last) | 1. Key information ☑<br>2. Balanced ☑ and informative ☑ | | |
| Text<br>(assess last) | 1. Sufficient detail others could reproduce ☑<br>2. Clear/concise writing ☑, table(s) ☑, diagram(s) ●, figure(s) ☑ | | |
| | | **Preliminaries [/5]** | 4 |
| **2. Introduction** | | | |
| Background | 1. Summary of current knowledge ☑<br>2. Specific problem(s) addressed ☑ and reason(s) for addressing ☑ | | |
| Objective | 1. Primary objective(s), hypothesis(es), or aim(s) ☑<br>2. Secondary question(s) ● | | |
| | **Is it worth continuing?** | **Introduction [/5]** | 5 |
| **3. Design** | | | |
| Research design | 1. Research design(s) chosen ☑ and why ☑<br>2. Suitability of research design(s) ☑ | | |
| Intervention, Treatment, Exposure | 1. Intervention(s)/treatment(s)/exposure(s) chosen ☑ and why ☑<br>2. Precise details of the intervention(s)/treatment(s)/exposure(s) ☑ for each group ☑<br>3. Intervention(s)/treatment(s)/exposure(s) valid ☑ and reliable ☑ | | |
| Outcome, Output, Predictor, Measure | 1. Outcome(s)/output(s)/predictor(s)/measure(s) chosen ☑ and why ☑<br>2. Clearly define outcome(s)/output(s)/predictor(s)/measure(s) ☑<br>3. Outcome(s)/output(s)/predictor(s)/measure(s) valid ☑ and reliable ☑ | | |
| Bias, etc | 1. Potential bias ☑, confounding variables ☒, effect modifiers ☒, interactions ☒<br>2. Sequence generation ●, group allocation ☑, group balance ☒, and by whom ●<br>3. Equivalent treatment of participants/cases/groups ☒ | different settings for each group's interview | |
| | **Is it worth continuing?** | **Design [/5]** | 4 |
| **4. Sampling** | | | |
| Sampling method | 1. Sampling method(s) chosen ☑ and why ☑<br>2. Suitability of sampling method ☑ | | |
| Sample size | 1. Sample size ☑, how chosen ☒, and why ☒<br>2. Suitability of sample size ☑ | | |
| Sampling protocol | 1. Target/actual/sample population(s): description ☑ and suitability ☑<br>2. Participants/cases/groups: inclusion ☑ and exclusion ☒ criteria<br>3. Recruitment of participants/cases/groups ☑ | | |
| | **Is it worth continuing?** | **Sampling [/5]** | 4 |
| **5. Data collection** | | | |
| Collection method | 1. Collection method(s) chosen ☑ and why ☑<br>2. Suitability of collection method(s) ☑ | | |
| Collection protocol | 1. Include date(s) ☑, location(s) ☑, setting(s) ☑, personnel ☑, materials ☑, processes ☑<br>2. Method(s) to ensure/enhance quality of measurement/instrumentation ☑<br>3. Manage non-participation ☑, withdrawal ☑, incomplete/lost data ● | | |
| | **Is it worth continuing?** | **Data collection [/5]** | 5 |
| **6. Ethical matters** | | | |
| Participant ethics | 1. Informed consent ☑, equity ☒<br>2. Privacy ☒, confidentiality/anonymity ☒ | | |
| Researcher ethics | 1. Ethical approval ☑, funding ☑, conflict(s) of interest ☑<br>2. Subjectivities ☑, relationship(s) with participants/cases ☒ | uncertain what relationship of researcher to providers are | |
| | **Is it worth continuing?** | **Ethical matters [/5]** | 3 |
| **7. Results** | | | |
| Analysis, Integration, Interpretation method | 1. A.I.I. method(s) for primary outcome(s)/output(s)/predictor(s) chosen ☑ and why ☑<br>2. Additional A.I.I. methods (e.g. subgroup analysis) chosen ● and why ●<br>3. Suitability of analysis/integration/interpretation method(s) ☑ | | |
| Essential analysis | 1. Flow of participants/cases/groups through each stage of research ☑<br>2. Demographic and other characteristics of participants/cases/groups ☑<br>3. Analyse raw data ☑, response rate ☑, non-participation/withdrawal/incomplete/lost data ☑ | | |
| Outcome, Output, Predictor analysis | 1. Summary of results ☑ and precision ☑ for each outcome/output/predictor/measure<br>2. Consideration of benefits/harms ☑, unexpected results ●, problems/failures ☑<br>3. Description of outlying data (e.g. diverse cases, adverse effects, minor themes) ☑ | | |
| | | **Results [/5]** | 5 |
| **8. Discussion** | | | |
| Interpretation | 1. Interpretation of results in the context of current evidence ☑ and objectives ☑<br>2. Draw inferences consistent with the strength of the data ☑<br>3. Consideration of alternative explanations for observed results ●<br>4. Account for bias ☑, confounding/effect modifiers/interactions/imprecision ☒ | | |
| Generalisation | 1. Consideration of overall practical usefulness of the study ☑<br>2. Description of generalisability (external validity) of the study ☑ | | |
| Concluding remarks | 1. Highlight study's particular strengths ☑<br>2. Suggest steps that may improve future results (e.g. limitations) ☑<br>3. Suggest further studies ☑ | | |
| | | **Discussion [/5]** | 4 |
| **9. Total** | | | |
| Total score | 1. Add all scores for categories 1–8 | | |
| | | **Total [/40]** | 34 |

### Crowe Critical Appraisal Tool (CCAT) Form (v1.4)

Reference: http://hdl.handle.net/10026.1/15469 Reviewer: CDM and ARC

This form must be used in conjunction with the CCAT User Guide (v1.4); otherwise validity and reliability may be severely compromised.

#### Citation

| Citation | Year |
|---|---|
| Using videoconsultations to deliver dietary advice to children with chronic kidney disease; a qualitative study of parent and child perspectives. | 2020 |

#### Research design (add if not listed)

| | |
|---|---|
| ❑ Not research | Article \| Editorial \| Report \| Opinion \| Guideline \| Pamphlet \| … |
| ❑ Historical | … |
| ☑ Qualitative | Narrative \| Phenomenology \| Ethnography \| Grounded theory \| Narrative case study \| … thematic analysis |
| ❑ Descriptive, Exploratory, Observational | A. Cross-sectional \| Longitudinal \| Retrospective \| Prospective \| Correlational \| Predictive \| … |
| | B. Cohort \| Case-control \| Survey \| Developmental \| Normative \| Case study \| … |
| Experimental: ❑ True experiment | Pre-test/post-test control group \| Solomon four-group \| Post-test only control group \| Randomised two-factor \| Placebo controlled trial \| … |
| Experimental: ❑ Quasi-experiment | Post-test only \| Non-equivalent control group \| Counter balanced *(cross-over)* \| Multiple time series \| Separate sample pre-test post-test [no Control] [Control] \| … |
| Experimental: ❑ Single system | One-shot experimental *(case study)* \| Simple time series \| One group pre-test/post-test \| Interactive \| Multiple baseline \| Within subjects *(Equivalent time, repeated measures, multiple treatment)* \| … |
| ❑ Mixed Methods | Action research \| Sequential \| Concurrent \| Transformative \| … |
| ❑ Synthesis | Systematic review \| Critical review \| Thematic synthesis \| Meta-ethnography \| Narrative synthesis \| … |
| ❑ Other | … |

#### Variables and analysis

| Intervention(s), Treatment(s), Exposure(s) | Outcome(s), Output(s), Predictor(s), Measure(s) | Data analysis method(s) |
|---|---|---|
| virtual consultation | patient perspectives | transcription of interviews verbatim, coding and charting of data, and thematic synthesis |

#### Sampling

| Total size | Group 1 | Group 2 | Group 3 | Group 4 | Control |
|---|---|---|---|---|---|
| 20 | 13 | 5 | 2 | | |

Population, sample, setting: Group 1 - 13 parents; Group 2 - 5 children; Group 3 - 2 dietitians

#### Data collection (add if not listed)

| | | | |
|---|---|---|---|
| Audit/Review | a) Primary \| Secondary \| …<br>b) Authoritative \| Partisan \| Antagonist \| …<br>c) Literature \| Systematic \| … | Interview | a) Formal \| Informal \| …<br>b) Structured \| Semi-structured \| Unstructured \| …<br>c) One-on-one \| Group \| Multiple \| Self-administered \| … |
| Observation | a) Participant \| Non-participant \| …<br>b) Structured \| Semi-structured \| Unstructured \| …<br>c) Covert \| Candid \| … | Testing | a) Standardised \| Norm-ref \| Criterion-ref \| Ipsative \| …<br>b) Objective \| Subjective \| …<br>c) One-on-one \| Group \| Self-administered \| … |

#### Scores

| | | | | | | | | | |
|---|---|---|---|---|---|---|---|---|---|
| Preliminaries | 5 | Design | 5 | Data Collection | 4 | Results | 4 | Total [/40] | 35 |
| Introduction | 5 | Sampling | 4 | Ethical Matters | 3 | Discussion | 5 | Total [%] | 87.5 |

#### General notes

CC BY-SA Crowe Critical Appraisal Tool (CCAT) :: Version 1.4 (19 November 2013) :: Michael Crowe
This work is licensed under the Creative Commons Attribution-ShareAlike 3.0 Unported License. To view a copy of this license, visit http://creativecommons.org/licenses/by-sa/3.0/

Appraise research on the merits of the research design used, not against other research designs.

| Category<br>Item | Item descriptors<br>[☑ Present; ☒ Absent; ■ Not applicable] | Description<br>[Important information for each item] | Score<br>[0–5] |
|---|---|---|---|
| **1. Preliminaries** | | | |
| Title | 1. Includes study aims ✓ and design ✓ | | |
| Abstract<br>(assess last) | 1. Key information ✓<br>2. Balanced ✓ and informative ✓ | | |
| Text<br>(assess last) | 1. Sufficient detail others could reproduce ✓<br>2. Clear/concise writing ✓, table(s) ❑, diagram(s) ❑, figure(s) ✓ | | |
| | | **Preliminaries [/5]** | 5 |
| **2. Introduction** | | | |
| Background | 1. Summary of current knowledge ✓<br>2. Specific problem(s) addressed ✓ and reason(s) for addressing ✓ | | |
| Objective | 1. Primary objective(s), hypothesis(es), or aim(s) ✓<br>2. Secondary question(s) ● | | |
| | **Is it worth continuing?** | **Introduction [/5]** | 5 |
| **3. Design** | | | |
| Research design | 1. Research design(s) chosen ✓ and why ✓<br>2. Suitability of research design(s) ✓ | | |
| Intervention, Treatment, Exposure | 1. Intervention(s)/treatment(s)/exposure(s) chosen ✓ and why ✓<br>2. Precise details of the intervention(s)/treatment(s)/exposure(s) ✓ for each group ●<br>3. Intervention(s)/treatment(s)/exposure(s) valid ✓ and reliable ✓ | | |
| Outcome, Output, Predictor, Measure | 1. Outcome(s)/output(s)/predictor(s)/measure(s) chosen ✓ and why ✓<br>2. Clearly define outcome(s)/output(s)/predictor(s)/measure(s) ✓<br>3. Outcome(s)/output(s)/predictor(s)/measure(s) valid ✓ and reliable ✓ | | |
| Bias, etc | 1. Potential bias ✓, confounding variables ✓, effect modifiers ☒, interactions ☒<br>2. Sequence generation ●, group allocation ●, group balance ●, and by whom ●<br>3. Equivalent treatment of participants/cases/groups ☒ | uncertain what the relationship of the researcher to participants is | |
| | **Is it worth continuing?** | **Design [/5]** | 5 |
| **4. Sampling** | | | |
| Sampling method | 1. Sampling method(s) chosen ✓ and why ✓<br>2. Suitability of sampling method ✓ | | |
| Sample size | 1. Sample size ✓, how chosen ✓, and why ✓<br>2. Suitability of sample size ✓ | | |
| Sampling protocol | 1. Target/actual/sample population(s): description ✓ and suitability ✓<br>2. Participants/cases/groups: inclusion ☒ and exclusion ☒ criteria<br>3. Recruitment of participants/cases/groups ✓ | | |
| | **Is it worth continuing?** | **Sampling [/5]** | 4 |
| **5. Data collection** | | | |
| Collection method | 1. Collection method(s) chosen ✓ and why ✓<br>2. Suitability of collection method(s) ✓ | | |
| Collection protocol | 1. Include date(s) ✓, location(s) ✓, setting(s) ✓, personnel ✓, materials ✓, processes ✓<br>2. Method(s) to ensure/enhance quality of measurement/instrumentation ✓<br>3. Manage non-participation ☒, withdrawal ☒, incomplete/lost data ☒ | | |
| | **Is it worth continuing?** | **Data collection [/5]** | 4 |
| **6. Ethical matters** | | | |
| Participant ethics | 1. Informed consent ☒, equity ✓<br>2. Privacy ☒, confidentiality/anonymity ☒ | | |
| Researcher ethics | 1. Ethical approval ✓, funding ✓, conflict(s) of interest ✓<br>2. Subjectivities ☒, relationship(s) with participants/cases ☒ | uncertain what relationship between researcher and participants is | |
| | **Is it worth continuing?** | **Ethical matters [/5]** | 3 |
| **7. Results** | | | |
| Analysis, Integration, Interpretation method | 1. A.I.I. method(s) for primary outcome(s)/output(s)/predictor(s) chosen ✓ and why ✓<br>2. Additional A.I.I. methods (e.g. subgroup analysis) chosen ● and why ●<br>3. Suitability of analysis/integration/interpretation method(s) ✓ | | |
| Essential analysis | 1. Flow of participants/cases/groups through each stage of research ✓<br>2. Demographic and other characteristics of participants/cases/groups ✓<br>3. Analyse raw data ✓, response rate ☒, non-participation/withdrawal/incomplete/lost data ☒ | | |
| Outcome, Output, Predictor analysis | 1. Summary of results ✓ and precision ● for each outcome/output/predictor/measure<br>2. Consideration of benefits/harms ✓, unexpected results ✓, problems/failures ✓<br>3. Description of outlying data (e.g. diverse cases, adverse effects, minor themes) ✓ | | |
| | | **Results [/5]** | 4 |
| **8. Discussion** | × | | |
| Interpretation | 1. Interpretation of results in the context of current evidence ✓ and objectives ✓<br>2. Draw inferences consistent with the strength of the data ✓<br>3. Consideration of alternative explanations for observed results ✓<br>4. Account for bias ✓, confounding/effect modifiers/interactions/imprecision ✓ | | |
| Generalisation | 1. Consideration of overall practical usefulness of the study ✓<br>2. Description of generalisability (external validity) of the study ✓ | | |
| Concluding remarks | 1. Highlight study's particular strengths ✓<br>2. Suggest steps that may improve future results (e.g. limitations) ✓<br>3. Suggest further studies ✓ | | |
| | | **Discussion [/5]** | 5 |
| **9. Total** | | | |
| Total score | 1. Add all scores for categories 1–8 | | |
| | | **Total [/40]** | 35 |

### Crowe Critical Appraisal Tool (CCAT) Form (v1.4)

Reference: https://doi.org/10.1186/s12882-021-02284-3 | Reviewer: CDM and ARC

This form must be used in conjunction with the CCAT User Guide (v1.4); otherwise validity and reliability may be severely compromised.

#### Citation

| Citation | Year |
|---|---|
| Video as an alternative to in-person consultations in outpatient renal transplant recipient follow-up: a qualitative study | 2021 |

#### Research design (add if not listed)

| | | |
|---|---|---|
| ❑ Not research | | Article \| Editorial \| Report \| Opinion \| Guideline \| Pamphlet \| … |
| ❑ Historical | | … |
| ☑ Qualitative | | Narrative \| Phenomenology \| Ethnography \| Grounded theory \| Narrative case study \| … thematic analysis |
| ❑ Descriptive, Exploratory, Observational | | A. Cross-sectional \| Longitudinal \| Retrospective \| Prospective \| Correlational \| Predictive \| … |
| | | B. Cohort \| Case-control \| Survey \| Developmental \| Normative \| Case study \| … |
| Experimental | ❑ True experiment | Pre-test/post-test control group \| Solomon four-group \| Post-test only control group \| Randomised two-factor \| Placebo controlled trial \| … |
| | ❑ Quasi-experiment | Post-test only \| Non-equivalent control group \| Counter balanced *(cross-over)* \| Multiple time series \| Separate sample pre-test post-test [no Control] [Control] \| … |
| | ❑ Single system | One-shot experimental *(case study)* \| Simple time series \| One group pre-test/post-test \| Interactive \| Multiple baseline \| Within subjects *(Equivalent time, repeated measures, multiple treatment)* \| … |
| ❑ Mixed Methods | | Action research \| Sequential \| Concurrent \| Transformative \| … |
| ❑ Synthesis | | Systematic review \| Critical review \| Thematic synthesis \| Meta-ethnography \| Narrative synthesis \| … |
| ❑ Other | | … |

#### Variables and analysis

| Intervention(s), Treatment(s), Exposure(s) | Outcome(s), Output(s), Predictor(s), Measure(s) | Data analysis method(s) |
|---|---|---|
| Patients using telemedicine vs in-person visits | patient perspectives on telemedicine compared to in-person | semi-structured interviews and thematic analysis of transcripts |

#### Sampling

| Total size | | Group 1 | | Group 2 | | Group 3 | | Group 4 | | Control |
|---|---|---|---|---|---|---|---|---|---|---|
| | 21 | | 15 | | 3 | | 3 | | | |

Population, sample, setting: 21 Participants were enrolled, with 15 patients (which I classify here as group 1), 3 providers (group 2), and 3 that did not participate in the post-study interviews (group 3)

#### Data collection (add if not listed)

| | | | |
|---|---|---|---|
| Audit/Review | a) Primary \| Secondary \| …<br>b) Authoritative \| Partisan \| Antagonist \| …<br>c) Literature \| Systematic \| … | Interview | a) Formal \| Informal \| …<br>b) Structured \| Semi-structured \| Unstructured \| …<br>c) One-on-one \| Group \| Multiple \| Self-administered \| … |
| Observation | a) Participant \| Non-participant \| …<br>b) Structured \| Semi-structured \| Unstructured \| …<br>c) Covert \| Candid \| … | Testing | a) Standardised \| Norm-ref \| Criterion-ref \| Ipsative \| …<br>b) Objective \| Subjective \| …<br>c) One-on-one \| Group \| Self-administered \| … |

#### Scores

| | | | | | | | | | |
|---|---|---|---|---|---|---|---|---|---|
| Preliminaries | 5 | Design | 4 | Data Collection | 5 | Results | 5 | Total [/40] | 36 |
| Introduction | 5 | Sampling | 3 | Ethical Matters | 4 | Discussion | 5 | Total [%] | 90 |

#### General notes

BY-SA This work is licensed under the Creative Commons Attribution-ShareAlike 3.0 Unported License. To view a copy of this license, visit http://creativecommons.org/licenses/by-sa/3.0/

Appraise research on the merits of the research design used, not against other research designs.

| Category<br>Item | Item descriptors<br>[☑ Present; ☒ Absent; ■ Not applicable] | Description<br>[Important information for each item] | Score<br>[0–5] |
|---|---|---|---|
| **1. Preliminaries** | | | |
| Title | 1. Includes study aims ☑ and design ☑ | | |
| Abstract<br>(assess last) | 1. Key information ☑<br>2. Balanced ☑ and informative ☑ | | |
| Text<br>(assess last) | 1. Sufficient detail others could reproduce ☑<br>2. Clear/concise writing ☑, table(s) ☑, diagram(s) ■, figure(s) ☑ | | |
| | | **Preliminaries [/5]** | 5 |
| **2. Introduction** | | | |
| Background | 1. Summary of current knowledge ☑<br>2. Specific problem(s) addressed ☑ and reason(s) for addressing ☑ | | |
| Objective | 1. Primary objective(s), hypothesis(es), or aim(s) ☑<br>2. Secondary question(s) ■ | | |
| | **Is it worth continuing?** Yes | **Introduction [/5]** | 5 |
| **3. Design** | | | |
| Research design | 1. Research design(s) chosen ☑ and why ☑<br>2. Suitability of research design(s) ☑ | | |
| Intervention, Treatment, Exposure | 1. Intervention(s)/treatment(s)/exposure(s) chosen ☑ and why ☒<br>2. Precise details of the intervention(s)/treatment(s)/exposure(s) ☑ for each group ■<br>3. Intervention(s)/treatment(s)/exposure(s) valid ☑ and reliable ☑ | | |
| Outcome, Output, Predictor, Measure | 1. Outcome(s)/output(s)/predictor(s)/measure(s) chosen ☑ and why ☑<br>2. Clearly define outcome(s)/output(s)/predictor(s)/measure(s) ☑<br>3. Outcome(s)/output(s)/predictor(s)/measure(s) valid ☑ and reliable ☑ | | |
| Bias, etc | 1. Potential bias ☑, confounding variables ☑, effect modifiers ☑, interactions ☑<br>2. Sequence generation ■, group allocation ■, group balance ■ and by whom ■<br>3. Equivalent treatment of participants/cases/groups ☒ | in limitations, the authors mention how patients were selected by nephrologist as stable and were all from 1 hopsital which could've affected the study | |
| | **Is it worth continuing?** | **Design [/5]** | 4 |
| **4. Sampling** | | | |
| Sampling method | 1. Sampling method(s) chosen ☑ and why ☑<br>2. Suitability of sampling method ☑ | | |
| Sample size | 1. Sample size ☑, how chosen ☒, and why ☒<br>2. Suitability of sample size ☑ | | |
| Sampling protocol | 1. Target/actual/sample population(s): description ☑ and suitability ☑<br>2. Participants/cases/groups: inclusion ☑ and exclusion ☑ criteria<br>3. Recruitment of participants/cases/groups ☑ | | |
| | **Is it worth continuing?** | **Sampling [/5]** | 3 |
| **5. Data collection** | | | |
| Collection method | 1. Collection method(s) chosen ☑ and why ☑<br>2. Suitability of collection method(s) ☑ | | |
| Collection protocol | 1. Include date(s) ☑, location(s) ☑, setting(s) ☑, personnel ☑, materials ☑, processes ☑<br>2. Method(s) to ensure/enhance quality of measurement/instrumentation ☑<br>3. Manage non-participation ☑, withdrawal ☒, incomplete/lost data ■ | | |
| | **Is it worth continuing?** | **Data collection [/5]** | 5 |
| **6. Ethical matters** | | | |
| Participant ethics | 1. Informed consent ☑, equity ☑<br>2. Privacy ☒, confidentiality/anonymity ☒ | all patients were telephone interviews whereas all providers were in-person interviews Privacy not mentioned for interiew |  |
| Researcher ethics | 1. Ethical approval ☑, funding ☑, conflict(s) of interest ☑<br>2. Subjectivities ☑, relationship(s) with participants/cases ☒ | uncertain what relationship of interviewers to interviewees is. Study not subjective as it was transcribed verbatim | |
| | **Is it worth continuing?** | **Ethical matters [/5]** | 4 |
| **7. Results** | | | |
| Analysis, Integration, Interpretation method | 1. A.I.I. method(s) for primary outcome(s)/output(s)/predictor(s) chosen ☑ and why ☑<br>2. Additional A.I.I. methods (e.g. subgroup analysis) chosen ■ and why ■<br>3. Suitability of analysis/integration/interpretation method(s) ☑ | | |
| Essential analysis | 1. Flow of participants/cases/groups through each stage of research ☑<br>2. Demographic and other characteristics of participants/cases/groups ☑<br>3. Analyse raw data ☑, response rate ☑, non-participation/withdrawal/incomplete/lost data ☑ | | |
| Outcome, Output, Predictor analysis | 1. Summary of results ☑ and precision ■ for each outcome/output/predictor/measure<br>2. Consideration of benefits/harms ☑, unexpected results ☑, problems/failures ☑<br>3. Description of outlying data (e.g. diverse cases, adverse effects, minor themes) ☑ | | |
| | | **Results [/5]** | 5 |
| **8. Discussion** | | | |
| Interpretation | 1. Interpretation of results in the context of current evidence ☑ and objectives ☑<br>2. Draw inferences consistent with the strength of the data ☑<br>3. Consideration of alternative explanations for observed results ☑<br>4. Account for bias ☑, confounding/effect modifiers/interactions/imprecision ☑ | | |
| Generalisation | 1. Consideration of overall practical usefulness of the study ☑<br>2. Description of generalisability (external validity) of the study ☑ | | |
| Concluding remarks | 1. Highlight study's particular strengths ☑<br>2. Suggest steps that may improve future results (e.g. limitations) ☑<br>3. Suggest further studies ☑ | | |
| | | **Discussion [/5]** | 5 |
| **9. Total** | | | |
| Total score | 1. Add all scores for categories 1–8 | | |
| | | **Total [/40]** | 36 |

#### Crowe Critical Appraisal Tool (CCAT) Form (v1.4)

Reference: https://www-clinicalkey-com.gcsom.idm.oclc.org/#!/content/playContent/1-s2.0-S221226721830995X?returnurl=null&referrer=null

Reviewer: CDM and ARC

This form must be used in conjunction with the CCAT User Guide (v1.4); otherwise validity and reliability may be severely compromised.

##### Citation

| Citation | Year |
|---|---|
| Patients' Experiences and Perspectives of Telehealth Coaching with a Dietitian to Improve Diet Quality in Chronic Kidney Disease: A Qualitative Interview Study | 2019 |

##### Research design (add if not listed)

| | | |
|---|---|---|
| ❏ Not research | | Article \| Editorial \| Report \| Opinion \| Guideline \| Pamphlet \| … |
| ❏ Historical | | … |
| ☑ Qualitative | | Narrative \| Phenomenology \| Ethnography \| Grounded theory \| Narrative case study \| … thematic analysis |
| ❏ Descriptive, Exploratory, Observational | | A. Cross-sectional \| Longitudinal \| Retrospective \| Prospective \| Correlational \| Predictive \| … |
| | | B. Cohort \| Case-control \| Survey \| Developmental \| Normative \| Case study \| … |
| Experimental | ❏ True experiment | Pre-test/post-test control group \| Solomon four-group \| Post-test only control group \| Randomised two-factor \| Placebo controlled trial \| … |
| | ❏ Quasi-experiment | Post-test only \| Non-equivalent control group \| Counter balanced *(cross-over)* \| Multiple time series \| Separate sample pre-test post-test [no Control] [Control] \| … |
| | ❏ Single system | One-shot experimental *(case study)* \| Simple time series \| One group pre-test/post-test \| Interactive \| Multiple baseline \| Within subjects *(Equivalent time, repeated measures, multiple treatment)* \| … |
| ❏ Mixed Methods | | Action research \| Sequential \| Concurrent \| Transformative \| … |
| ❏ Synthesis | | Systematic review \| Critical review \| Thematic synthesis \| Meta-ethnography \| Narrative synthesis \| … |
| ❏ Other | | … |

##### Variables and analysis

| Intervention(s), Treatment(s), Exposure(s) | Outcome(s), Output(s), Predictor(s), Measure(s) | Data analysis method(s) |
|---|---|---|
| Dietary telehealth using telephone calls and text messages | Patient demographics and perspectives on intervention | Transcription of the interviews which were then coded on HyperRESEARCH followed by theme development |

##### Sampling

| Total size | | Group 1 | | Group 2 | | Group 3 | | Group 4 | | Control |
|---|---|---|---|---|---|---|---|---|---|---|
| | 21 | | | | | | | | | |

Population, sample, setting: patients aged 28-78 with stages 3-4 of chronic kidney disease

##### Data collection (add if not listed)

| | | | |
|---|---|---|---|
| Audit/Review | a) Primary \| Secondary \| …<br>b) Authoritative \| Partisan \| Antagonist \| …<br>c) Literature \| Systematic \| … | Interview | a) Formal \| Informal \| …<br>b) Structured \| Semi-structured \| Unstructured \| …<br>c) One-on-one \| Group \| Multiple \| Self-administered \| … |
| Observation | a) Participant \| Non-participant \| …<br>b) Structured \| Semi-structured \| Unstructured \| …<br>c) Covert \| Candid \| … | Testing | a) Standardised \| Norm-ref \| Criterion-ref \| Ipsative \| …<br>b) Objective \| Subjective \| …<br>c) One-on-one \| Group \| Self-administered \| … |

##### Scores

| | | | | | | | | | |
|---|---|---|---|---|---|---|---|---|---|
| Preliminaries | 5 | Design | 4 | Data Collection | 5 | Results | 5 | Total [/40] | 38 |
| Introduction | 5 | Sampling | 5 | Ethical Matters | 4 | Discussion | 5 | Total [%] | 95 |

##### General notes

CC BY-SA
Crowe Critical Appraisal Tool (CCAT) :: Version 1.4 (19 November 2013) :: Michael Crowe
This work is licensed under the Creative Commons Attribution-ShareAlike 3.0 Unported License. To view a copy of this license, visit http://creativecommons.org/licenses/by-sa/3.0/

Appraise research on the merits of the research design used, not against other research designs.

| **Category**<br>Item | **Item descriptors**<br>[☑ Present; ☒ Absent; ■ Not applicable] | **Description**<br>**[Important information for each item]** | **Score**<br>[0–5] |
|---|---|---|---|
| **1. Preliminaries** | | | |
| Title | 1. Includes study aims ☑ and design ☑ | | |
| Abstract<br>(assess last) | 1. Key information ☑<br>2. Balanced ☑ and informative ☑ | | |
| Text<br>(assess last) | 1. Sufficient detail others could reproduce ☑<br>2. Clear/concise writing ☑, table(s) ☑, diagram(s) ■, figure(s) ☑ | | |
| | | **Preliminaries [/5]** | 5 |
| **2. Introduction** | | | |
| Background | 1. Summary of current knowledge ☑<br>2. Specific problem(s) addressed ☑ and reason(s) for addressing ☑ | | |
| Objective | 1. Primary objective(s), hypothesis(es), or aim(s) ☑<br>2. Secondary question(s) ■ | | |
| | **Is it worth continuing?** Yes | **Introduction [/5]** | 5 |
| **3. Design** | | | |
| Research design | 1. Research design(s) chosen ☑ and why ☑<br>2. Suitability of research design(s) ☑ | | |
| Intervention, Treatment, Exposure | 1. Intervention(s)/treatment(s)/exposure(s) chosen ☑ and why ☑<br>2. Precise details of the intervention(s)/treatment(s)/exposure(s) ☑ for each group ■<br>3. Intervention(s)/treatment(s)/exposure(s) valid ☑ and reliable ☑ | reason for intervention found at end of intro | |
| Outcome, Output, Predictor, Measure | 1. Outcome(s)/output(s)/predictor(s)/measure(s) chosen ☑ and why ☑<br>2. Clearly define outcome(s)/output(s)/predictor(s)/measure(s) ☑<br>3. Outcome(s)/output(s)/predictor(s)/measure(s) valid ☑ and reliable ☑ | | |
| Bias, etc | 1. Potential bias ☑, confounding variables ☒, effect modifiers ☒, interactions ☒<br>2. Sequence generation ❑, group allocation ■, group balance ■, and by whom ■<br>3. Equivalent treatment of participants/cases/groups ☒ | one participant had face-to-face interview whereas all others were by phone | |
| | **Is it worth continuing?** | **Design [/5]** | 4 |
| **4. Sampling** | | | |
| Sampling method | 1. Sampling method(s) chosen ☑ and why ☒<br>2. Suitability of sampling method ☑ | | |
| Sample size | 1. Sample size ☑, how chosen ☑, and why ☑<br>2. Suitability of sample size ☑ | participants added until saturation | |
| Sampling protocol | 1. Target/actual/sample population(s): description ☑ and suitability ☑<br>2. Participants/cases/groups: inclusion ☑ and exclusion ☒ criteria<br>3. Recruitment of participants/cases/groups ☑ | | |
| | **Is it worth continuing?** Yes | **Sampling [/5]** | 5 |
| **5. Data collection** | | | |
| Collection method | 1. Collection method(s) chosen ☑ and why ☑<br>2. Suitability of collection method(s) ☑ | | |
| Collection protocol | 1. Include date(s) ☑, location(s) ☒, setting(s) ☑, personnel ☑, materials ☑, processes ☑<br>2. Method(s) to ensure/enhance quality of measurement/instrumentation ☑<br>3. Manage non-participation ☑, withdrawal ☒, incomplete/lost data ☒ | only 1 person approached did not agree and was not included in the study | |
| | **Is it worth continuing?** | **Data collection [/5]** | 5 |
| **6. Ethical matters** | | | |
| Participant ethics | 1. Informed consent ☑, equity ☑<br>2. Privacy ☒, confidentiality/anonymity ☒ | privacy not explicitly mentioned | |
| Researcher ethics | 1. Ethical approval ☑, funding ☑, conflict(s) of interest ☑<br>2. Subjectivities ☑, relationship(s) with participants/cases ☑ | | |
| | **Is it worth continuing?** | **Ethical matters [/5]** | 4 |
| **7. Results** | | | |
| Analysis, Integration, Interpretation method | 1. A.I.I. method(s) for primary outcome(s)/output(s)/predictor(s) chosen ☑ and why ☑<br>2. Additional A.I.I. methods (e.g. subgroup analysis) chosen ■ and why ■<br>3. Suitability of analysis/integration/interpretation method(s) ☑ | | |
| Essential analysis | 1. Flow of participants/cases/groups through each stage of research ☑<br>2. Demographic and other characteristics of participants/cases/groups ☑<br>3. Analyse raw data ☑, response rate ☑, non-participation/withdrawal/incomplete/lost data ☑ | | |
| Outcome, Output, Predictor analysis | 1. Summary of results ☑ and precision ■ for each outcome/output/predictor/measure<br>2. Consideration of benefits/harms ☑, unexpected results ☒, problems/failures ■<br>3. Description of outlying data (e.g. diverse cases, adverse effects, minor themes) ☑ | | |
| | | **Results [/5]** | 5 |
| **8. Discussion** | | | |
| Interpretation | 1. Interpretation of results in the context of current evidence ☑ and objectives ☑<br>2. Draw inferences consistent with the strength of the data ☑<br>3. Consideration of alternative explanations for observed results ■<br>4. Account for bias ☑, confounding/effect modifiers/interactions/imprecision ☒ | | |
| Generalisation | 1. Consideration of overall practical usefulness of the study ☑<br>2. Description of generalisability (external validity) of the study ☑ | | |
| Concluding remarks | 1. Highlight study's particular strengths ☑<br>2. Suggest steps that may improve future results (e.g. limitations) ☑<br>3. Suggest further studies ☑ | | |
| | | **Discussion [/5]** | 5 |
| **9. Total** | | | |
| Total score | 1. Add all scores for categories 1–8 | | |
| | | **Total [/40]** | 38 |
